# Cell-type-specific transcriptome architecture underlying the establishment and exacerbation of systemic lupus erythematosus

**DOI:** 10.1101/2022.01.12.22269137

**Authors:** Masahiro Nakano, Mineto Ota, Yusuke Takeshima, Yukiko Iwasaki, Hiroaki Hatano, Yasuo Nagafuchi, Takahiro Itamiya, Junko Maeda, Ryochi Yoshida, Saeko Yamada, Aya Nishiwaki, Haruka Takahashi, Hideyuki Takahashi, Yuko Akutsu, Takeshi Kusuda, Hiroyuki Suetsugu, Lu Liu, Kwangwoo Kim, Xianyong Yin, So-Young Bang, Yong Cui, Hye-Soon Lee, Hirofumi Shoda, Xuejun Zhang, Sang-Cheol Bae, Chikashi Terao, Kazuhiko Yamamoto, Tomohisa Okamura, Kazuyoshi Ishigaki, Keishi Fujio

## Abstract

Systemic lupus erythematosus (SLE) is a complex and heterogeneous autoimmune disease involving multiple immune cells. A major hurdle to the elucidation of SLE pathogenesis is our limited understanding of dysregulated gene expression linked to various clinical statuses with a high cellular resolution. Here, we conducted a large-scale transcriptome study with 6,386 RNA sequencing data covering 27 immune cell types from 159 SLE and 89 healthy donors. We first profiled two distinct cell-type-specific transcriptomic signatures: disease-state and disease-activity signatures, reflecting disease establishment and exacerbation, respectively. We next identified candidate biological processes unique to each signature. This study suggested the clinical value of disease-activity signatures, which were associated with organ involvement and responses to therapeutic agents such as belimumab. However, disease-activity signatures were less enriched around SLE risk variants than disease-state signatures, suggesting that the genetic studies to date may not well capture clinically vital biology in SLE. Together, we identified comprehensive gene signatures of SLE, which will provide essential foundations for future genomic, genetic, and clinical studies.

## Introduction

Systemic lupus erythematosus (SLE) is a systemic autoimmune disease that involves multiple immune cell types and pathways^1,2^. SLE has a broad spectrum of clinical manifestations such as skin rashes, arthritis and nephritis, and the disease course is generally unpredictable^3^. This heterogeneous nature has hampered a better understanding of SLE pathogenesis and the development of effective therapeutic agents^4,5^. To date, only two biologics have been approved for SLE, belimumab (BLM) and anifrolumab, monoclonal antibodies against B cell-activating factor (BAFF) and type I interferon (IFN) receptor subunit 1, respectively^6–9^.

To detect biomarkers and therapeutic targets for SLE, several studies on bulk whole-blood or peripheral blood mononuclear cell (PBMC) transcriptomes have revealed some key gene signatures related to IFN signaling, granulocytes, and plasma cells^10–16^. However, these studies have suffered from one critical limitation: the results were biased by the abundance of various immune cell populations in the analyzed samples, which complicates the identification of any cell-type-specific disease-relevant signatures^17^. Therefore, recent studies applied single-cell RNA sequencing (scRNA-seq), a powerful technology to improve cellular resolution, to PBMCs, skin and kidney samples from SLE patients and have successfully identified several cell subpopulations crucial for lupus pathogenesis^18–20^. However, since these scRNA-seq studies of SLE were limited by sparse expression information and small sample sizes (around 30 cases), they were not well-powered to capture comprehensive transcriptome abnormality related to different clinical manifestations. These limitations could be overcome by a large-scale bulk transcriptome study with finely sorted cell populations.

SLE etiology has both genetic and environmental components^1–3^. Researchers have conducted large-scale genome-wide association studies (GWASs) for SLE^21–23^, identifying more than one hundred risk loci. Combined with omics data mostly from healthy individuals, researchers attempted to interpret the genetic etiology and have identified potential causal roles of IFN, Toll-like receptor signaling and immune complexes^24–26^. However, these studies have not thoroughly investigated the complex interactions between risk variants and the transcriptome dysregulation seen in SLE patients. Such investigations hold the promise to elucidate the complex pathogenesis of SLE.

To address these issues, we conducted a large-scale transcriptome study of 6,386 bulk RNA sequencing (RNA-seq) data including 27 purified immune cell types in peripheral blood that encompassed almost every type of immune cell (**Fig. 1**). We recruited 136 SLE patients with various disease activities and clinical presentations (22 among them were re-evaluated after BLM treatment; **Methods**) and 89 healthy volunteers in the Immune Cell Gene Expression Atlas from the University of Tokyo (ImmuNexUT) cohort^27^ (discovery dataset). Using multiple approaches, we investigated cell-type-specific transcriptome dysregulation and classified them into two categories: disease-state and disease-activity signatures. Furthermore, we deployed these signatures to five main topics of downstream analyses: i) replication, ii) biological interpretation, iii) diverse organ involvement, iv) pre- and post-treatment comparison, and v) the SLE-GWAS signals (**Fig. 1**). Overall, our large-scale and comprehensive investigation uncovered the molecular basis underlying the clinical heterogeneity of SLE with a fine resolution of cell-type-specificity.

**Fig. 1.**
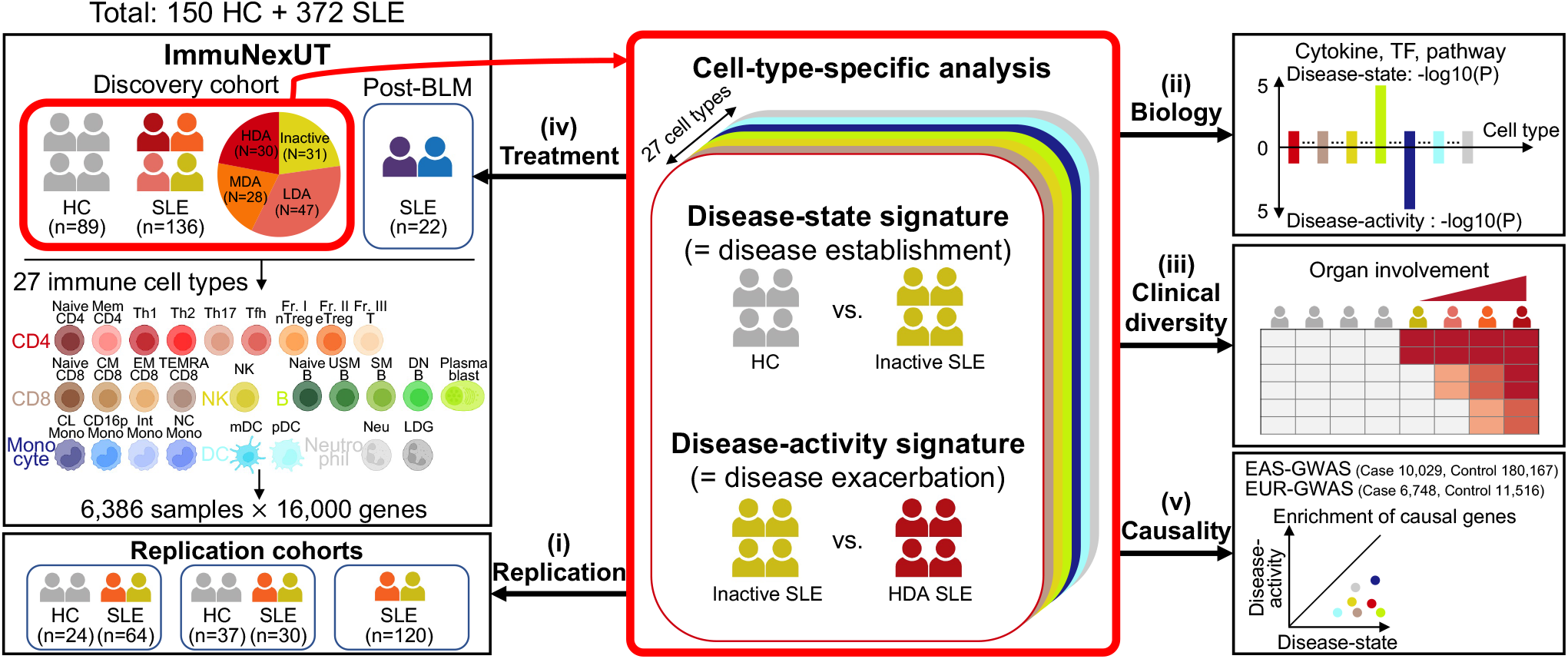
Overview of this study. We profiled 6,386 RNA sequencing data of 27 immune cell types from peripheral blood in HC and SLE patients **(left)**. We identified two distinct categories of disease-relevant signatures in a cell-type-specific manner **(middle)**, and then performed extensive downstream analyses **(i-v)**. BLM, belimumab; EAS, East Asian; EUR, European; TF, transcription factor.

## Results

### Overview of gene expression patterns in the ImmuNexUT cohort

Our dataset included 27 finely sorted immune cell types: CD4^+^ T cells, nine subsets; CD8^+^ T cells, four subsets; NK cells, one subset; B cells, five subsets; monocytes, four subsets; dendritic cells, two subsets; and neutrophils, two subsets (**Fig. 1, left**; **Supplementary Table 1**). We recruited 248 donors in total. Among them, 136 unique SLE patients and 89 healthy controls (HC) were included in the discovery dataset; the rest included 22 post-BLM patients (**Methods**). Compared with previous studies with fine resolution transcriptomes^18–20^, larger sample size with multiple clinical statuses (e.g., disease activity, organ involvement, and treatment profiles) is an advantage of our cohort (**Supplementary Note**; **Supplementary Table 2**). At enrollment, 30 patients (22.1%) in the discovery dataset had high disease activity (HDA; SLEDAI-2K^28^ ≥ 9), while 31 (22.8%) patients were inactive (SLEDAI-2K = 0). Forty one (30.1%), 27 (19.9%) and 30 (22.1%) patients had mucocutaneous, musculoskeletal, and renal activity, respectively^29^.

To understand the highly complex transcriptomic signatures in our datasets (16,000 genes and 6,386 samples from 248 donors; **Extended Data Fig. 1a**; **Methods**), we first aimed to project all samples in low dimensional spaces. Principal component analysis (PCA) showed that the samples from the same cell type clustered together followed by the same cell lineage (**Extended Data Fig. 1b**); this pattern became more evident in the uniform manifold approximation and projection (UMAP) (**Extended Data Fig. 1c**). We also confirmed that batch effects were successfully removed in all cell types (**Extended Data Fig. 2a-b**).

IFN-related gene (IRG) expression is a hallmark signature of SLE^30–32^. To explicitly quantify transcriptome patterns well-established for lupus, we utilized 100 IRGs reported in a recent PBMC scRNA-seq study having the highest cellular resolution^20^ (**Fig. 2a**). The cell-type- and disease-specific IRG expression patterns were globally consistent with those in the original publication: e.g., upregulated *CXCL10* and *IFITM3* expression (G5) in lupus CD16-positive monocytes (CD16p Mono) and upregulated *IRF7* and *PARP10* expression (G8) in plasmacytoid dendritic cells (pDC). Additionally, about half (n=54) of IRGs showed the highest expression in SLE neutrophil-lineage cells, which were not evaluated in the previous scRNA-seq studies of SLE^18–20^. Together, our transcriptomic data exhibited the expected cell-type-specific patterns and recapitulated previously established IRG signatures.

**Fig. 2.**
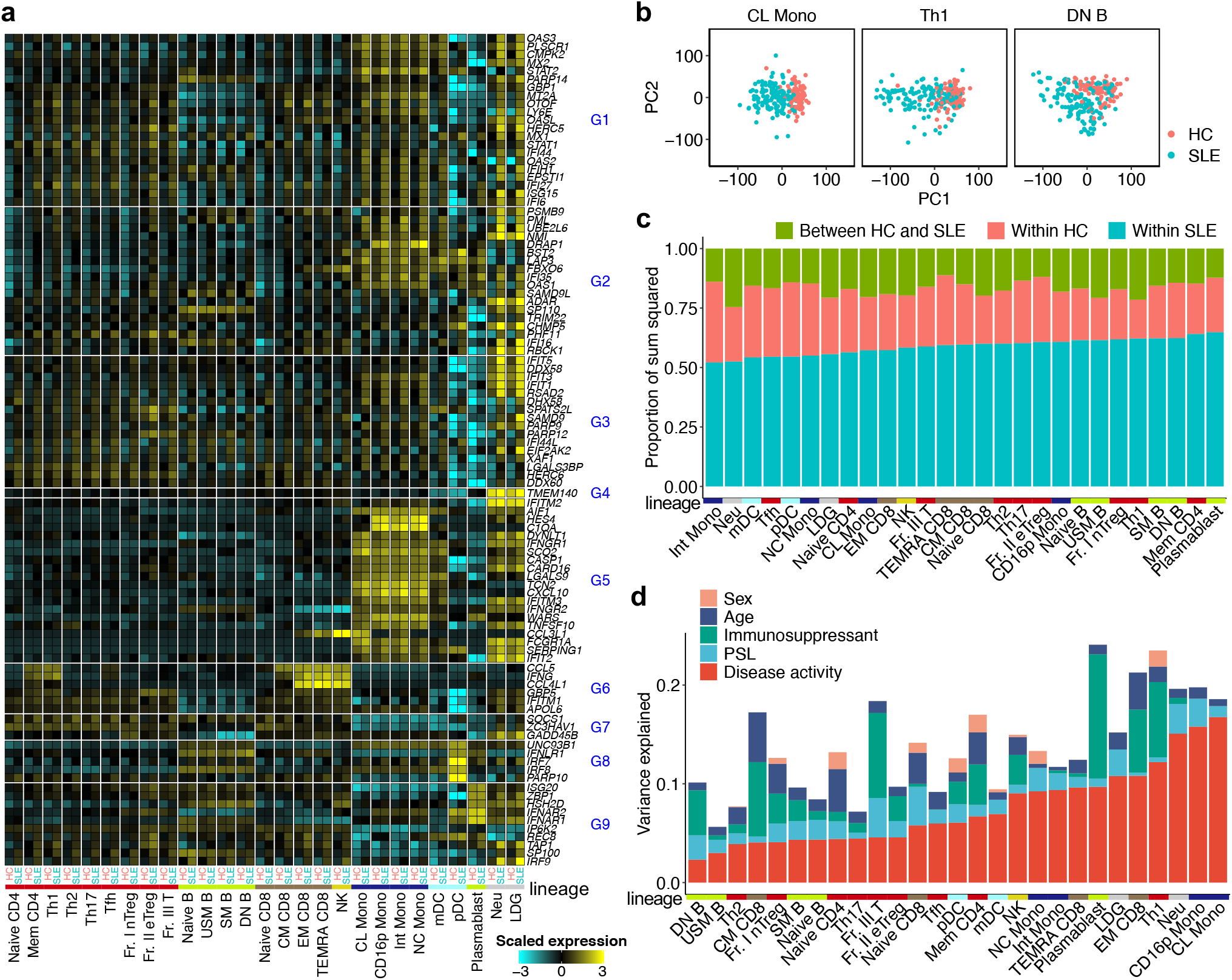
Overview of gene expression patterns in the ImmuNexUT cohort. **a,** A heatmap showing the mean expression levels of 100 IRGs across all 27 cell types and diseases. Expression levels are scaled for each gene. Genes and clusters (G1-9) originate from a PBMC scRNA-seq study^20^. Cell types are arranged consistently with the original publication where applicable. **b,** PCA plots of HC and SLE gene expression data in representative cell types (see also **Extended Data Fig. 3**). **c,** A bar plot showing the proportion of sum squared deviations within HC, SLE, and between HC and SLE data in each cell type. Cell types are arranged based on the sum squared deviations within SLE. **d,** A bar plot showing the proportion of variance explained by the clinical parameters within SLE data in each cell type. Cell types are arranged based on the variance explained by disease activity. In **a** and **c-d,** column annotation colors indicate cell lineages. We used the discovery dataset (n=225) for all analyses in this figure. PSL, prednisolone.

### Disease activity is a major source of variation in the within-SLE transcriptome data

To explore the source of the transcriptomic variations in the discovery dataset, we conducted PCA within each cell type and evaluated the distribution of samples in the PCA space (**Supplementary Data 1**). The top PCs differentiated SLE patients from HC in all cell types, indicating widespread transcriptome perturbations in SLE immune cells (**Fig. 2b; Extended Data Fig. 3**). In addition, gene expression profiles within patients showed higher variation than those within HC (**Fig. 2c; Methods**).

We further evaluated how the transcriptome variations reflected the heterogeneous clinical statuses. We first confirmed that PC1-7 is the minimum set to associate the transcriptome with the clinical parameters in the discovery dataset and utilized PC1-7 scores for subsequent analyses (**Supplementary Note**; **Supplementary Fig. 1a-c; Supplementary Table 3**). We then quantified the contribution of clinical parameters to the within-SLE variation using weighted variance partitioning analysis (**Fig. 2d; Methods**). Importantly, this analysis revealed that disease activity had the largest contribution to the total variation within SLE in almost all cell types (7.6% on average), around 2.9-fold larger than the treatment contribution.

### SLE disease-state and activity signatures

Motivated by the fact that both case-control differences and disease activity substantially contributed to the whole transcriptome architecture, we next deployed a supervised approach to the discovery dataset to identify two transcriptomic signatures for each cell type: i) disease-state signature genes, defined as differentially expressed genes (DEGs, false discovery rate [FDR] < 0.05) between inactive SLE and HC, which reflect the biology of disease establishment, and ii) disease-activity signature genes, defined as DEGs between HDA and inactive SLE, which reflect the biology of disease exacerbation (**Fig. 1, middle; Fig. 3a; Extended Data Fig. 4a**). We detected comparable numbers of DEGs between these two comparisons (on average, 2,098 disease-state and 2,114 disease-activity signature genes; **Extended Data Fig. 4b; Supplementary Data 2-3**). We conducted replication analysis using independent cohorts^33–35^ (**Fig. 1, i**) and confirmed the robustness of both signatures by showing their high replicability (one-sided sign test, Bonferroni-corrected *P* < 0.05; **Supplementary Note; Supplementary Table 4; Extended Data Fig. 5a-b**).

**Fig. 3.**
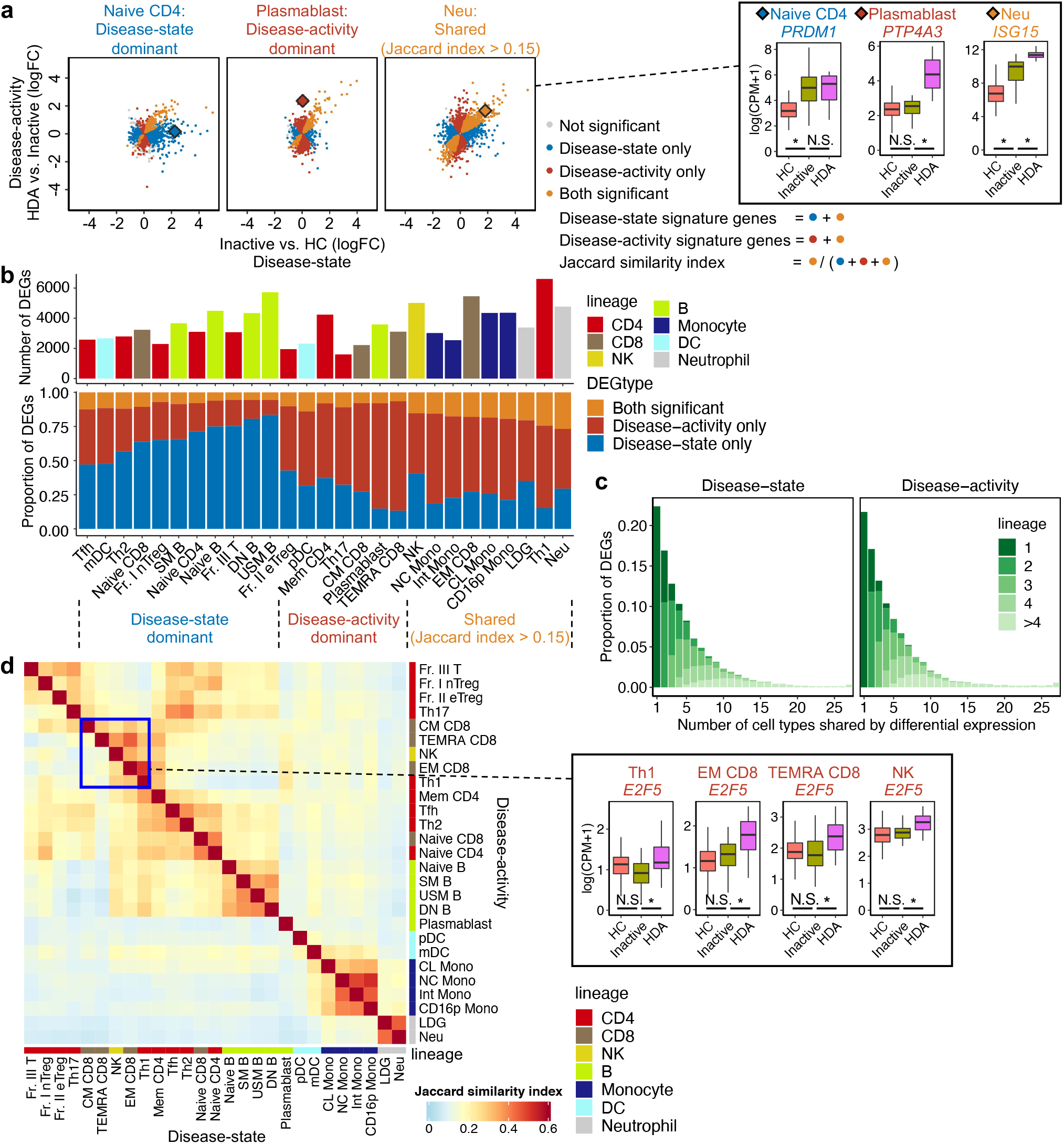
SLE disease-state and activity signatures. **a, (left)** Scatter plots comparing the logFC of disease-state and activity signature genes in representative cell types (see also **Extended Data Fig. 4a**). Colors indicate the significance of each signature**. (right)** Box plots showing the expression of representative disease-state, activity and both significant signature genes. **b,** Bar plots showing **(top)** the number of the union of disease-state/activity signature genes and **(bottom)** the proportion of DEG types in each cell type. Cell types are separated into three groups (**Methods**). **c,** Histograms showing the proportion of the number of cell types sharing DEG for both signatures. Colors indicate the number of shared cell lineages. **d, (left)** A heatmap showing the Jaccard similarity indexes across all cell types in both signatures. The order of cell types in row and column are same and based on the hierarchical clustering using the Jaccard indexes of disease-state signatures. **(right)** Box plots showing the expression of a representative disease-activity signature gene shared by Th1, NK, and CD8+ memory T-lineage cells. Within each boxplot in **b** and **d**, the horizontal lines reflect the median, the top and bottom of each box reflect the interquartile range (IQR), and the whiskers reflect the maximum and minimum values within each grouping no further than 1.5 x IQR from the hinge. *, DEG (FDR < 0.05); N.S., not significant. We used the discovery dataset (n=225) for all analyses in this figure.

To examine the specificity of these signatures, we first compared them within each cell type. We calculated the Jaccard similarity index in each cell type to quantify the shared genes between signatures (**Fig. 3a**); we considered that a gene was shared when it was included in both signatures with a concordant sign (**Methods**). Based on the proportion of DEGs and Jaccard index, we found three different transcriptome perturbation patterns (**Methods**): disease-state dominant pattern, e.g., B-lineage cells and naive CD4/8+ T cells, disease-activity dominant pattern, e.g., plasmablasts, and shared pattern, e.g., monocyte- and neutrophil-lineage cells (**Fig. 3a-b; Extended Data Fig. 4b**).

When we evaluated the similarity based on the correlation of log fold changes (logFC) for both signatures, we observed consistent patterns (**Extended Data Fig. 4c**). We next compared signature genes across different cell types and confirmed cell-type-specific and shared components. Around 20% and 30% of signature genes were detected in only one cell type and lineage, respectively (**Fig. 3c**). To understand the distribution of the shared components across cell types, we calculated the Jaccard index (**Fig. 3d**) and correlation (**Extended Data Fig. 4d**). Globally, we detected higher similarity within the same cell-lineage than in different lineages in both signatures. However, we found a clear discrepancy between the signatures; high similarity among T helper 1 (Th1) and cytotoxic lymphocytes (natural killer [NK] and CD8+ memory T-lineage cells)^36^ was observed only in activity signatures (blue square in **Fig. 3d** and **Extended Data Fig. 4d**). These gaps indicated the presence of gene expression patterns specific to HDA patients in these cell types. Thus, our dataset captured distinct transcriptome perturbations in the disease establishment and exacerbation phases in a cell-type-specific manner.

### Cell-type-specific biology in disease establishment and exacerbation

To understand the SLE biology in disease establishment and exacerbation, we next sought to interpret disease-state and activity signature genes in each cell type using multiple external databases (**Fig. 1, ii**).

First, we focused on 137 genes encoding cytokines (**Supplementary Table 5**), the key regulators of immune responses and potential drug targets in autoimmune diseases^37^. Fifty-one genes were upregulated in at least one signature, consistent with previous studies: *IFNG* in Th1, NK, and CD8+ memory T-lineage cells^38,39^, and *TNFSF13B*, encoding BAFF, especially in DC-, monocyte-, and neutrophil-lineage cells with the highest expression^40^ (**Fig. 4a, top**). Among these genes, we identified 21 and 17 that were upregulated specifically in one cell-lineage in disease-state and activity signatures, respectively; representative examples of activity signatures included *IL12A/B* in switched memory B cells (SM B), *IL1B* in monocyte-lineage, *CCL2/8* in classical monocyte (CL mono), *IL18/TNFSF15* in neutrophil-lineage cells, and *IL21* and *CXCL13* in Th1 (**Fig. 4a, top**). Among them, *IL21* and *CXCL13*, critical genes to support B cell antibody production^41,42^, are especially intriguing. Although previous studies reported follicular helper T cells (Tfh) as the major source of *IL21* and *CXCL13*^43^, Th1 showed a more dynamic increase than Tfh, especially in the activity signatures (**Fig. 4a, bottom**).

**Fig. 4.**
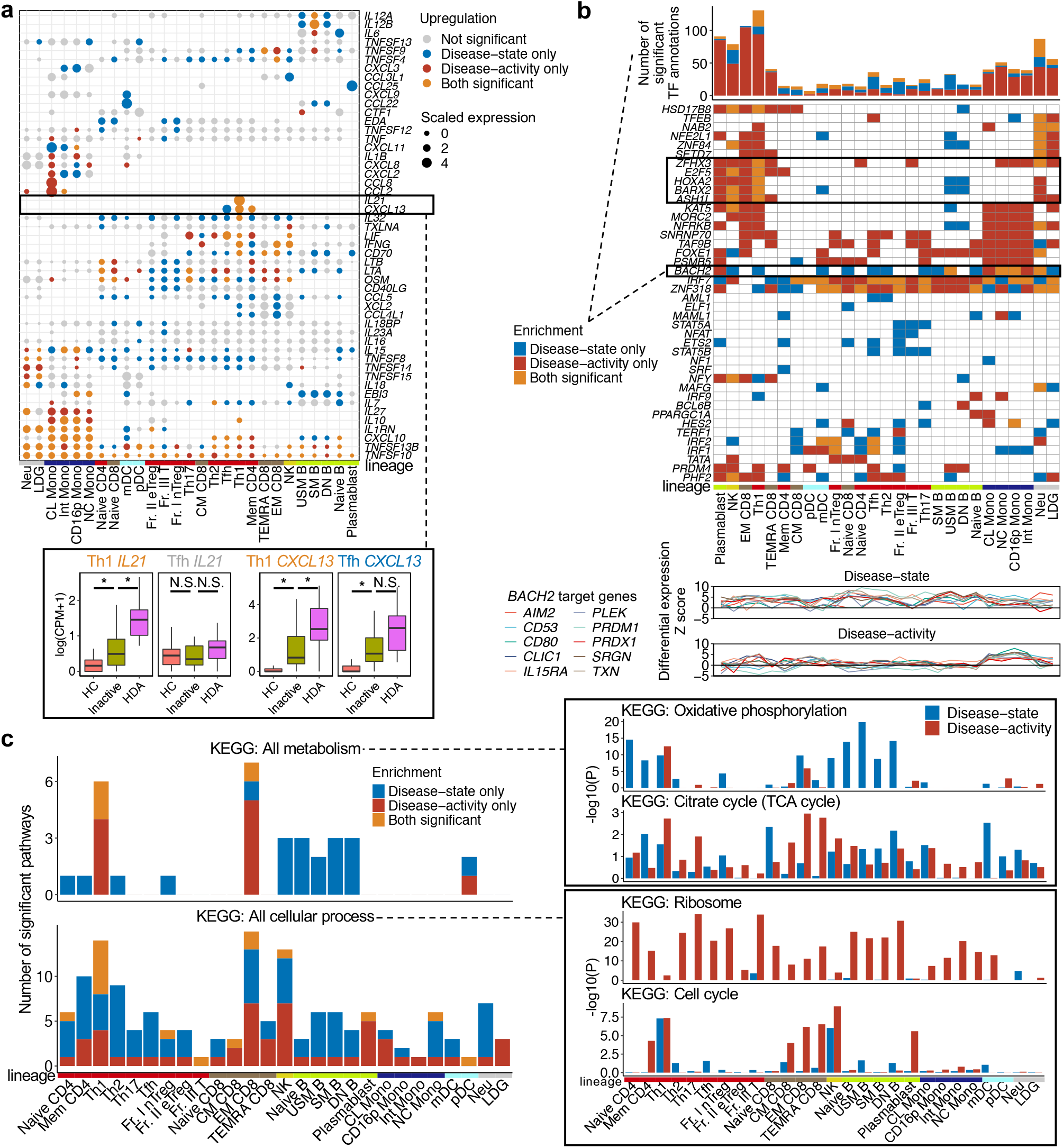
Cell-type-specific biology in disease establishment and exacerbation. **a, (top)** Upregulated cytokines as disease-state and/or activity signature genes for each cell type. Genes and cell types are hierarchically clustered based on differential expression Z scores of activity signatures. **(bottom)** Boxplots showing the expression in representative cytokines. The horizontal lines reflect the median, the top and bottom of each box reflect the IQR, and the whiskers reflect the maximum and minimum values within each grouping no further than 1.5 x IQR from the hinge. *, DEG (FDR < 0.05); N.S., not significant. **b, (top)** A bar plot showing the number of significant TF annotation enrichments for each signature. **(middle)** A heatmap showing TF enrichment for each signature. TFs and cell types are hierarchically clustered based on –log10(enrichment *P*) of activity signatures. Only the top three TFs with strongest enrichments in each signature are shown, excluding redundant annotations. **(bottom)** Line graphs showing the differential expression Z scores of 10 representative *BACH2* target genes in each cell type for both signatures. **c,** Bar plots showing **(left)** the number of significant enrichments of metabolism- and cellular process-related pathways, and **(right)** the enrichment of representative pathways for each signature. *P*, *P* values in one-sided Fisher’s exact test. We used the discovery dataset (n=225) for all analyses in this figure.

Next, we inferred activities of transcription factors (TF), essential regulators of immune function, based on the expression of TF-downstream genes (**Methods**). Among 61,182 total tests (1,133 TF annotations × 27 cell types × two signatures), we observed 1,228 significant enrichments for 299 annotations (FDR < 0.05; one-sided Fisher’s exact test; **Supplementary Table 6**). Among them, disease-activity signatures showed more enrichments (862 enrichments [70.2%]; **Fig. 4b, top**). These results suggested underappreciated pathogenic roles of TFs in SLE exacerbation. We here highlight two such examples (**Fig. 4b, middle**). First, cell cycle regulators including E2F-families showed strong enrichment in activity signatures of Th1, NK, CD8+ memory T-lineage cells, and plasmablasts, indicating that these cells are probably proliferating in active SLE patients. Upregulated cell cycle regulation might be driving the high similarities between these cell types in disease-activity signatures (**Fig. 3d, right**). Second, *BACH2* showed significant enrichment in disease-activity signatures for myeloid-lineage cells. Intriguingly, *BACH2* also showed strong enrichment in lymphocytes, consistent with previous studies^44,45^, but primarily in disease-state signatures. These results demonstrated that an identical gene regulatory machinery can exert a pathogenic effect in different cell types depending on the disease phases (**Fig. 4b, bottom**).

Lastly, we also performed pathway enrichment analyses to examine multiple biological processes underlying lupus pathogenesis (**Methods**). Among 32,292 total tests for 598 pathways, we observed 735 and 315 significant enrichments for disease-state and activity, respectively (FDR < 0.05; one-sided Fisher’s exact test; **Extended Data Fig. 6a; Supplementary Table 7-8**). We confirmed the enrichment of established pathways such as complement activation^46,47^ (**Supplementary Note; Extended Data Fig. 6b**). Intriguingly, we found different enrichment patterns between the signatures in metabolism- and cellular process-related KEGG pathways (**Fig. 4c, left**; **Extended Data Fig. 6c; Methods**). For example, oxidative phosphorylation signaling was enriched especially in B-lineage cells for disease-state whereas it was enriched in Th1 and effector memory CD8+ T cells (EMCD8) for disease-activity signatures (**Fig. 4c, right**). TCA cycle signaling was enriched in disease-activity signatures of Th1 and CD8+ memory T-lineage cells. Ribosome pathways were enriched only in disease-activity signatures. Cell cycle activation was enriched predominantly in disease-activity signatures of Th1, NK, CD8+ memory T-lineage cells, and plasmablasts. Although ribosome and cell cycle pathways were already described as disease activity-related pathways in previous bulk whole-blood studies^13,16^, our analysis clarified the precise cell-type origin of these pathways. Furthermore, we extended our view to previously underappreciated pathways such as immunometabolism, describing disease establishment and exacerbation phases separately.

### Cell-type-specific contribution to organ involvement in SLE

To resolve the complex relationships between transcriptome dysregulation and clinical heterogeneity in SLE, we leveraged a PC-based unsupervised approach (**Fig. 1, iii**). In the hierarchical clustering of 225 unique individuals using all 189 PCs (= 7 PCs × 27 cell types), HC were clearly separated from patients; in addition, HDA patients with multiple organ complications were clustered together (**Fig. 5a; Extended Data Fig. 7a**).

**Fig. 5.**
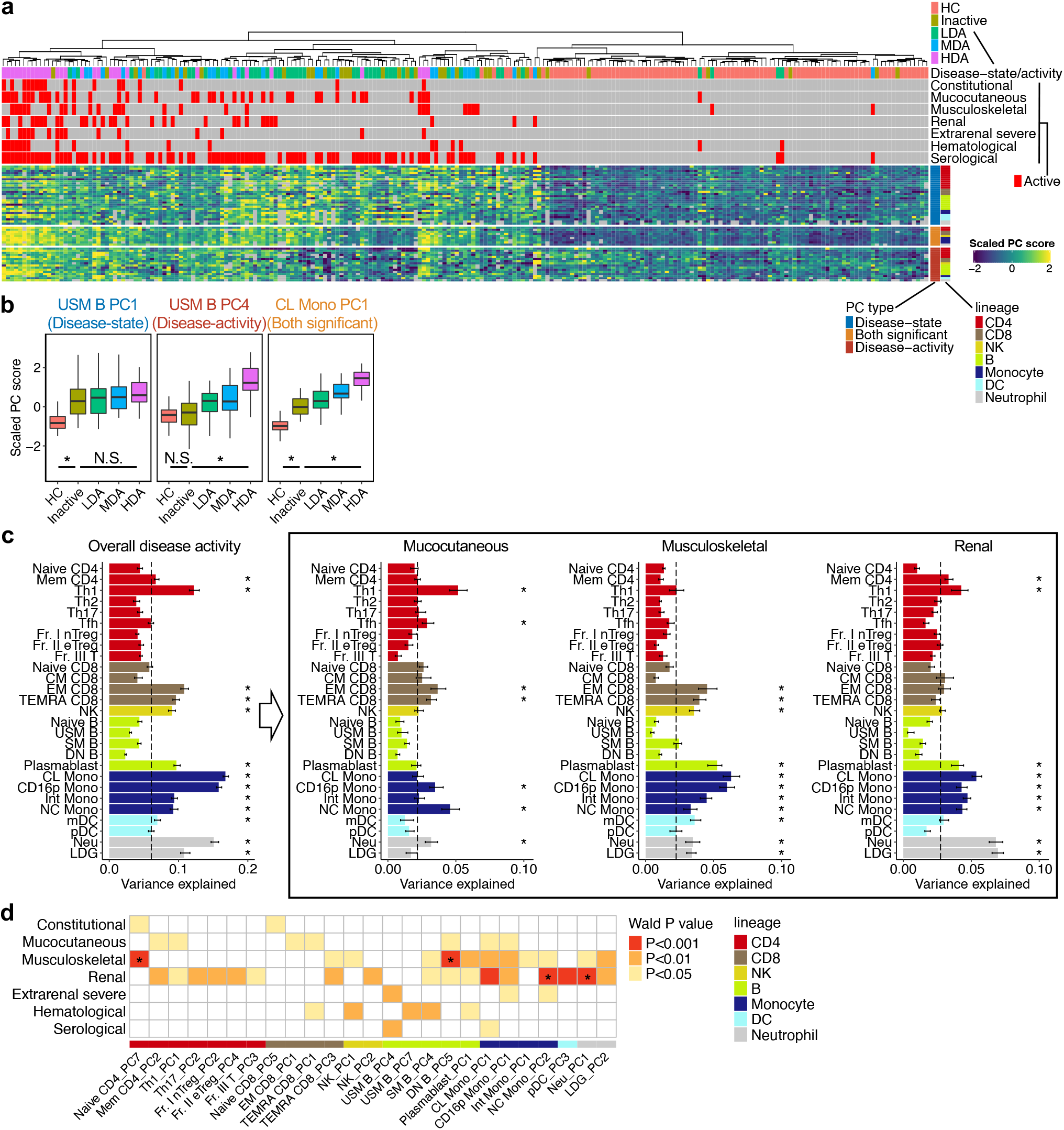
Cell-type-specific contribution to organ involvement in SLE. **a,** Hierarchical clustering of 225 unique individuals based on all PC1-7 scores of 27 cell types. Top annotations indicate the disease status and organ/domain activities in each individual. Right annotations indicate the type and cell lineage of each PC. Here, only disease-state and/or activity PCs are shown in the heatmap (see also **Extended Data Fig. 7a**). **b,** Box plots showing the scaled PC scores in representative disease-state, activity and both significant PCs. The horizontal lines reflect the median, the top and bottom of each box reflect the IQR, and the whiskers reflect the maximum and minimum values within each grouping no further than 1.5 x IQR from the hinge. *, FDR < 0.05 in linear regression test; N.S., not significant. **c,** Bar plots showing the proportion of variance explained by **(left)** the overall disease activity and **(right)** representative organ/domain activities within SLE data in each cell type (see also **Extended Data Fig. 8**). Error bars and dashed vertical lines indicate 95% confidence intervals from jackknife resampling and the median values across 27 cell types, respectively. *, Bonferroni-adjusted *P*jk < 0.05 (**Methods**). **d,** A heatmap showing the association of disease-activity PCs and organ/domain activities in SLE. *P*, nominal *P* values; *, FDR < 0.05 in linear regression test. We used the discovery dataset (n=225)

Compared with the approach using thousands of signature genes, the PC-based approach is better at representing whole transcriptome architecture with a small number of parameters. To understand the biological significance of each PC, we defined two categories of PCs as in the discussion of signature genes: i) disease-state PCs, separating inactive SLE and HC, and ii) disease-activity PCs, separating HDA and inactive SLE (FDR < 0.05; **Fig. 5b; Supplementary Table 3; Methods**). Among 189 PCs, we identified 37 disease-state PCs and 25 disease-activity PCs; among them, nine PCs were classified into both (**Extended Data Fig. 7b**). When we projected the data from independent cohorts onto our PCA space, PC scores maintained the original contrasts, confirming the good replicability of both PC signatures (one-sided sign test, *P* <0.05; **Supplementary Note; Extended Data Fig. 5c; Supplementary Table 9**). This PC-based approach successfully captured the continuous nature of SLE biology; most disease-activity PCs showed a gradual increase in the association signals along with the extent of disease activity (**Fig. 5b; Extended Data Fig. 7b**).

To overview cell-type-specific contributions to organ involvement, we first assessed the variance proportion of cell-type-specific PCs explained by clinical parameters (weighted variance partitioning analysis; **Methods**). Overall disease activity, a composite measure reflecting the status of all organs, significantly contributed to the within-SLE transcriptome variation especially in the 13 cell types including Th1, plasmablasts, and monocyte- and neutrophil-lineage cells (Bonferroni-corrected jackknife resampling *P* [*P_jk_*] < 0.05; **Fig. 2d; Fig. 5c, left; Methods**). We then decomposed the overall activity into seven organ/domain categories^28,29^: constitutional, mucocutaneous, musculoskeletal, renal, extrarenal severe, hematological, and serological activities (**Fig 5c, right; Extended Data Fig. 8; Supplementary Table 10; Methods**). Interestingly, each organ/domain showed distinct cell-type-specific patterns. While Th1 showed the highest explained variance for mucocutaneous activity, the contribution of monocyte-lineage cells was predominant for musculoskeletal activity. Furthermore, neutrophil-lineage cells exhibited the largest contribution to renal involvement, followed by monocyte-lineage cells, Th1, and plasmablasts.

We next evaluated the specific relationship of each disease-activity PC with organ involvement (**Fig 5d; Supplementary Note; Supplementary Table 3**). For renal activity, neutrophil (Neu) PC1 and non-classical monocyte (NC Mono) PC2 showed strong associations (linear regression test; FDR < 0.05). We also identified significant associations of Naive CD4 PC7 and double negative (DN) B cell PC5 with musculoskeletal activity; these associations might be underestimated by the weighted variance partitioning analysis, which prioritizes the contribution of top PCs (**Methods**). Together, our results confirmed the critical roles of granulocytes and macrophages for the development of lupus nephritis (LN)^48,49^. In addition, our results also suggested other potential cell-type-specific contributions to organ involvement, which may be informative in unravelling SLE clinical heterogeneity.

### Cell-type-specific activity signatures linked to treatment responses

Belimumab (BLM) is a monoclonal antibody that inhibits BAFF, a vital factor for B cell survival and differentiation^6,7,40^. We investigated the effect of BLM on the transcriptome in each cell type (**Fig. 1, iv**). Our cohort has longitudinal data before and six months after BLM induction on 22 individuals; we refer to them as pre- and post-BLM. Importantly, none of the post-BLM samples were included in the discovery dataset; therefore, the comparison between pre- and post-BLM is independent of the disease-activity signatures calculated in the discovery dataset. We observed DEGs between pre- and post-BLM (BLM-DEGs) predominantly in B-lineage cells, confirming the cell-type-specific effects of BLM (**Fig. 6a**). When we categorized the patients into good and poor responders to BLM (n = 9 and 13, respectively; **Methods**), more DEGs were observed in good than in poor responders (**Fig. 6b; Supplementary Data 4**). The IFNγ-related, nuclear factor-kappa B (NFκB)-related, and glycolysis-related pathways were enriched in B cell DEGs of good responders (FDR < 0.05, one-sided Fisher’s exact test; **Fig. 6c; Supplementary Table 11**), consistent with the downstream signaling of BAFF-receptors in B cells^50,51^.

**Fig. 6.**
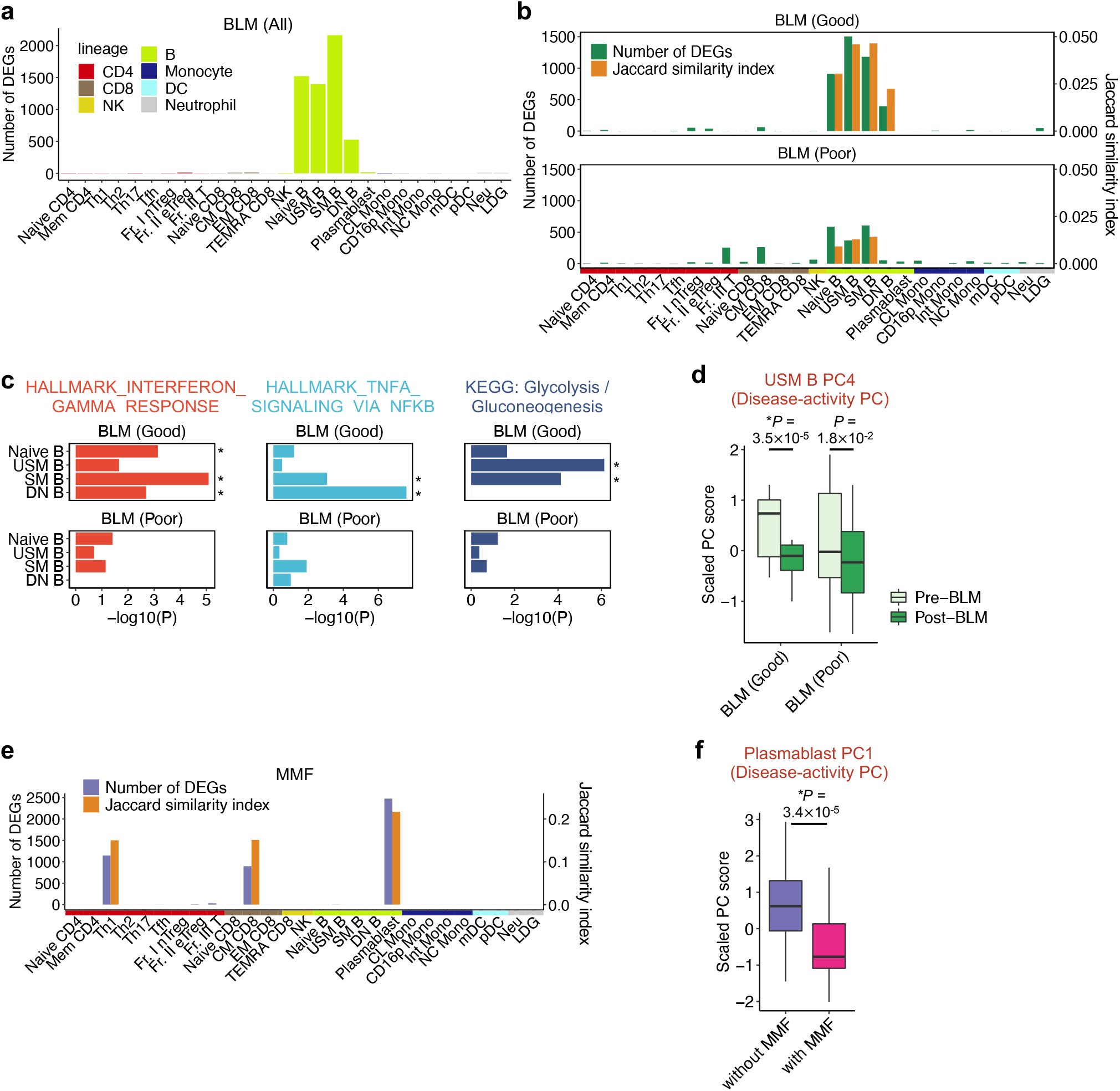
Cell-type-specific activity signatures linked to treatment responses. **a,** A bar plot showing the numbers of DEGs in each cell type between all post- vs. pre-BLM patients (n=22 paired samples). **b,** Bar plots showing the numbers of BLM-DEGs and Jaccard similarity indexes between BLM-DEGs and disease-activity signature genes in each cell type, separated into good (n=9) and poor (n=13) responders. **c,** Bar plots showing the enrichment of representative pathways for the BLM-DEGs in B-lineage cells, separated into good and poor responders. *P*, nominal *P* values; *, FDR < 0.05 in one-sided Fisher’s exact test. **d,** A box plot showing the USM B PC 4 scores from pre- and post-BLM, separated into good and poor responders. *P*, nominal *P* values; *, FDR < 0.05 in linear mixed regression test. **e,** Bar plots showing the numbers of DEGs between patients with (n=31) and without (n=105) MMF (MMF-DEGs), and Jaccard indexes between MMF-DEGs and disease-activity signature genes in each cell type. **f,** A box plot showing the plasmablasts PC 1 scores from patients with or without MMF. *P*, nominal *P* values; *, FDR < 0.05 in linear regression test. Within each boxplot in **d** and **f**, the horizontal lines reflect the median, the top and bottom of each box reflect the IQR, and the whiskers reflect the maximum and minimum values within each grouping no further than 1.5 x IQR from the hinge. In **b** and **e**, column annotation colors indicate cell lineages.

We next asked whether BLM effects on transcriptomes counteract disease-activity signatures. We first calculated the Jaccard similarity index to quantify the shared genes between BLM-DEGs and disease-activity signatures; we considered a gene is shared when the BLM effect had the opposite sign to the activity signature to reflect therapeutic responses (**Method**). Jaccard indexes in good responders were around 3.4-fold higher than those in poor responders in B-lineage cells (**Fig. 6b**); the analyses based on logFC also showed similar trends. (**Supplementary Note; Extended Data Fig. 9a-b**). By projecting post-BLM data onto the PCA space of the discovery cohort (**Methods**), we evaluated the change in disease-activity PC scores between pre- and post-BLM. Consistent with the DEG analysis, we found a significant decrease in unswitched memory B cells (USM B) PC4 scores only for good responders (linear mixed regression test, FDR = 9.9×10^-3^; **Fig. 6d**). Together, these results provided robust evidence supporting the association between disease-activity signatures and the therapeutic response to BLM.

Next, we assessed the effect of other therapeutic agents such as mycophenolate mofetil (MMF) on disease-activity signatures (**Supplementary Data 5; Supplementary Fig. 2**). Importantly, we treated disease activity as a potential confounder in this analysis (**Methods**), and hence the MMF effects are not biased by disease activity. DEGs between patients with and without MMF (MMF-DEGs) were primarily observed in plasmablasts, followed by Th1 and central memory CD8+ T cells (CM CD8) (**Fig. 6e**); the same cell types were nominated by variance partitioning analysis (**Extended Data Fig. 9c; Supplementary Table 10**). MMF-DEGs were shared with the activity signature genes with 15.0-21.7% of the Jaccard index (**Fig. 6e**). Pathway analysis recapitulated this shared component: both genes showed enrichment for oxidative phosphorylation and E2F-related cell cycle pathways (**Extended Data Fig. 9d; Supplementary Table 12**). We also found a significant decrease in disease-activity PC scores of plasmablasts (PC1) in patients taking MMF, adjusted for disease activity (linear regression test, FDR = 6.3×10^-3^; **Fig. 6f; Methods**). These results were consistent with previous reports that MMF suppressed plasma cell differentiation^52,53^.

### Risks variants for SLE are enriched around disease-state signatures, not activity signatures

To estimate the causal roles of these signatures for the risk of disease onset, we integrated our transcriptome data with the results of GWAS for SLE (SLE-GWAS; **Fig. 1, v**).

First, we analyzed the genome-wide distribution of all risk variants irrespective of their effect sizes and tested their enrichment around the signature genes using stratified linkage disequilibrium score regression^26,54^ (S-LDSC; **Methods**). To be in line with previous studies of S-LDSC, we additionally took a conventional approach and analyzed the enrichment of specifically expressed genes (SEG) in each cell type derived from HC (HC-SEG; **Methods**). Since ancestry-specific GWAS results are required in S-LDSC, we used two large-scale SLE-GWAS conducted in East Asian (EAS)^23^ and European (EUR)^22^ populations. We confirmed that the S-LDSC results for EAS-and EUR-GWAS were globally similar as reported for other traits^55^ (r in enrichment estimate = 0.69; *P* = 1.5×10^-12^; **Extended Data Fig. 10a; Supplementary Table 13**); hence we combined them using a fixed-effect meta-analysis. Consistent with previous reports^26^, the risk variants were predominantly enriched around HC-SEG of B-lineage cells but only with a nominal significance (minimum *P* = 0.028 for SM B): no significant enrichment at Bonferroni-corrected *P* < 0.05 (**Fig. 7a**). Strikingly, compared to HC-SEG, we found much stronger enrichments for disease-state signatures in all cell types: nine significant enrichments. However, for disease-activity signatures, we found no significant enrichment. Consistently, the enrichments were substantially weaker in the activity signatures than in the disease-state signatures (paired Wilcoxon test, *P* = 5.5×10^-6^).

**Fig. 7.**
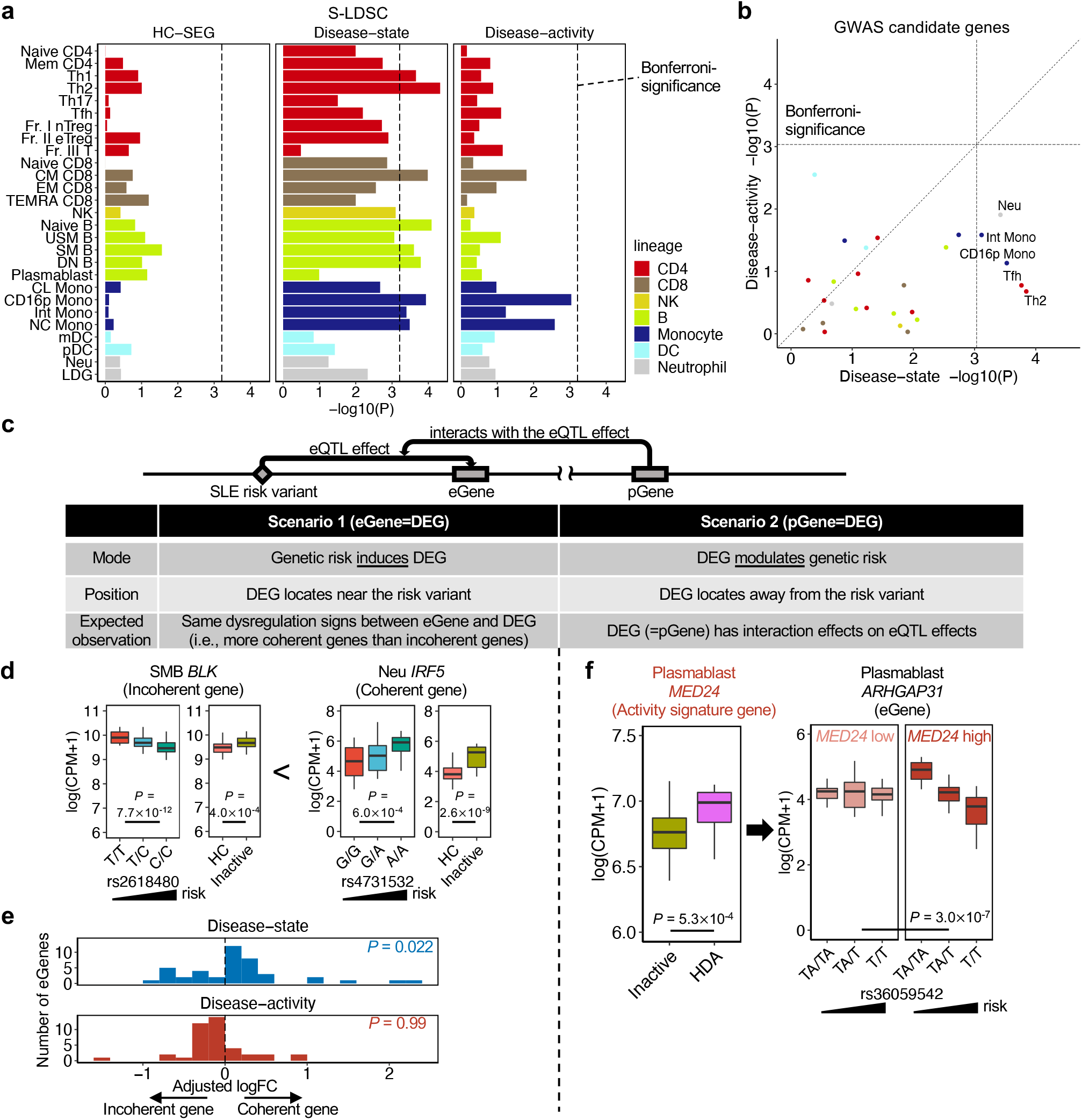
Risks variants for SLE are enriched around disease-state signatures, not activity signatures. **a,** Bar plots showing the enrichment of SLE risk variants around HC-SEG, disease-state and activity signatures for each cell type. *P*, enrichment *P* values in S-LDSC. Vertical dashed lines represent Bonferroni-significance. **b,** A scatter plot comparing the enrichment of SLE-GWAS candidate genes in disease-state and activity signatures for each cell type. Only the cell types that pass Bonferroni-significance (dashed lines) are annotated. *P*, *P* values in one-sided Fisher’s exact test. **c,** Hypothesis of the association between the risk variants and DEGs in SLE via eQTL effect. **d,** Box plots showing the expression patterns in representative coherent and incoherent genes. *P*, *P* values in linear regression (left) and differential expression test (right). **e,** Histogram of adjusted logFC in coherent and incoherent genes for disease-state and activity signatures. *P*, *P* values in one-sided sign test. **f,** Box plots showing **(left)** the differential expression of *MED24*, an activity signature gene in plasmablasts and **(right)** the influence of *MED24* on the eQTL effect of a SLE risk variant rs36059542 on *ARHGAP31*. *P*, *P* values in differential expression (left) and ANOVA test (right). Within each boxplot in **d** and **f**, the horizontal lines reflect the median, the top and bottom of each box reflect the IQR, and the whiskers reflect the maximum and minimum values within each grouping no further than 1.5 x IQR from the hinge. We used the discovery dataset (n=225) for all analyses in this figure.

We next analyzed candidate causal genes implicated in SLE-GWAS (**Methods; Supplementary Table 14**) and tested their enrichment in both signatures of each cell type (**Fig. 7b; Extended Data Fig. 10b; Supplementary Table 15**). As in the S-LDSC results, the enrichments of the candidate genes were predominantly observed in disease-state rather than in activity signatures; significant enrichments were found in five and zero cell types, respectively (Bonferroni-corrected *P* < 0.05). Again, the enrichments were weaker in activity signatures than disease-state signatures (paired Wilcoxon test, *P* = 2.5×10^-3^). These results confirmed that the risk variants locate on average around disease-state and not activity signature genes.

Considering that most of the risk variants are within the gene regulatory regions^25^, we hypothesized that risk variants possess gene regulatory effects on disease-state genes (i.e., eQTL effects; **Fig. 7c**). To test this hypothesis, we next evaluated the potential eQTL effects of the risk variants on disease-state and activity signature genes. Specifically, we focused on genes affected by the risk variants’ eQTL effects (eGenes) and asked how the risk alleles’ directional effects on eGenes are consistent with our gene expression signatures; an eGene with consistent direction is called a “coherent gene”^56^ (**Fig. 7d**; for this analysis, we utilized the colocalization test results between SLE-GWAS risk variants and eQTL variants that we recently reported^27^). Intriguingly, the coherent genes were enriched in disease-state signatures, but not in activity signatures (**Fig. 7e; Supplementary Table 16**): 67% of eGene-cell type combinations were coherent in disease-state signatures whereas only 25% of combinations were coherent in activity signatures (one-sided sign test, *P* = 0.022 and 0.99, respectively). Together, these analyses demonstrated better directional compatibility of the SLE risk allele’s effect with disease-state signatures rather than activity signatures. Although this might be reasonable considering most GWAS are based on case-control design, this finding implied the failure of current GWAS to capture the critical biology of SLE represented by disease-activity signatures.

While activity signature genes did not locate around the current SLE risk variants, we speculated that some of them might contribute to the disease risk by modulating the eQTL effects of risk alleles (we refer to the genes with modulating eQTL effects as proxy genes [pGenes]^57^; **Fig. 7c**). Therefore, we finally sought to test whether disease-activity signature genes act as pGenes for risk alleles in SLE patients. We included 115 patients with available genotyping data and examined the influence of disease-activity signatures genes on the eQTL effects of risk alleles (we again utilized the abovementioned colocalization results^27^). Intriguingly, we detected two significant pGenes among activity signature genes (ANOVA test, FDR < 0.05; **Supplementary Data 6**), which included *MED24,* a transcriptional coactivator. *MED24* is an activity signature gene of plasmablasts (**Fig. 7f, left**), and its expression was suppressed by MMF (**Extended Data Fig. 10c**). *MED24* significantly modulated the eQTL effect of a SLE risk variant (rs36059542) on *ARHGAP31,* encoding a GTPase-activating protein (ANOVA test, FDR = 0.035; **Fig. 7f, right**). Of note, the eQTL effect of rs36059542 on *ARHGAP31* was observed only in plasmablasts^27^. Therefore, in addition to disease-state genes, disease-activity genes may also contribute to genetic risk by modulating the eQTL effects of risk alleles. Furthermore, MMF might indirectly suppress SLE genetic risk by controlling the *MED24* expression.

## Discussion

In this study, we extensively examined dysregulated gene expression patterns of SLE by profiling 27 immune cell types from 89 HC and 159 SLE donors. We identified two distinct categories of lupus-relevant signatures: disease-state and activity genes. These signatures revealed multiple novel and underappreciated mechanisms with a fine cellular resolution. Moreover, we demonstrated the value of these signatures in multiple applications: transferability to independent transcriptome datasets, shared components with drug responses and consistency with the genetic signals.

SLE is a chronic disease characterized by relapsing and remitting disease course^1–3^. A critical but unsolved question is how the multi-cellular pathophysiology is different (or not different) between disease development and exacerbation stages. Most of the previous SLE transcriptome studies have failed to provide clear answers since they separately conducted case-control and/or intra-case analyses and have not directly compared both signatures in the same study with a high cellular resolution. Our comprehensive transcriptome data successfully clarified the multiple cell-type-specific immune-mediated pathways characteristic of disease-state and activity, which highlighted the distinct biology behind the establishment and exacerbation phases of this disease.

The current realistic goal of SLE management is to achieve remission or low disease activity, not a cure^58–60^. From this perspective, the disease-activity signature, not the disease-state signature, has implications for development of biomarkers of treatment response or therapeutic targets. Indeed, we observed shared components between the disease-activity signatures and transcriptome changes in BLM good responders and in those taking MMF. The cell-type-specific activity signatures identified in this study, e.g., myeloid-lineage cells characteristic of renal disease, might be informative for designing novel treatment strategies for SLE.

We have provided in-depth integrative analyses of the risk variants and transcriptome signatures; these results have several important implications. First, since we recruited established SLE patients in the chronic phase, whether the disease-state signature genes indicate causality (the signatures induce disease-state) or a reverse-causality (the disease-state induces the signatures) is a critical question. Since the risk variants reflect the causality, the fact that the risk variants are enriched around disease-state signature genes suggested the former scenario, further supported by the consistency in the dysregulation direction (i.e., coherent genes). Moreover, this finding also suggested that genetic risk-driven susceptibility signatures remained in clinically stable SLE patients, indicating that the causal mechanism is not completely controlled by the current treatments. Second, the current GWAS signals failed to reflect the disease-activity signatures. This is a critical limitation of the current genetic studies considering the potential importance of activity signatures in drug target discovery. To resolve this issue, a new framework of genetic study focusing on intra-case heterogeneity (e.g., disease severity) will be required.

Although our study has substantially improved our understanding of SLE biology, we need to acknowledge several limitations. First, apart from the pre- and post-BLM sub-cohort, our study lacked longitudinal data. Second, all participants in this study were from the EAS population, although we demonstrated the applicability of our signatures to transcriptome data and GWAS results from multiple ancestries. Third, our cell-sorting strategy might have failed to characterize currently unidentified cell populations.

It is now clear that disease-state and activity signatures jointly maintain the complex pathophysiology of SLE. These signatures have the potential to be a pivotal foundation for future genomic, genetic, and drug discovery studies.

## Methods

### Subjects

The data in our study was generated by the ImmuNexUT consortium^27^, approved by the Ethics Committees of the University of Tokyo. All participants were from the EAS ancestry. Healthy volunteers were recruited at the Department of Allergy and Rheumatology at the University of Tokyo Hospital. SLE patients were recruited at the Department of Allergy and Rheumatology at the University of Tokyo Hospital, Division of Rheumatic Diseases at National Center for Global Health and Medicine, or Immuno-Rheumatology Center at St. Luke’s International Hospital. Written informed consent was obtained from all participants. We have complied with all of the relevant ethical regulations.

All SLE patients met the 1997 revised version of the American College of Rheumatology classification criteria^61^. The exclusion criteria for the discovery dataset (including 136 SLE patients) were: i) active malignancies or infections, ii) use of more than 20mg prednisolone (PSL) daily or equivalent at enrollment, iii) receive of intravenous methylprednisolone pulse, cyclophosphamide, rituximab, or BLM within 12 months before enrollment.

In addition, we also collected 22 paired samples just before and six months after the additional therapy of BLM (i.e., pre- and post-BLM). Among the pre-BLM samples, 21 samples were included in the discovery dataset, and one was re-sampled due to the interval between initial enrollment and BLM induction. None of the post-BLM samples were included in the discovery dataset. Therefore, we recruited 248 donors in total: 136 unique SLE patients, one pre-BLM, 22 post-BLM patients, and 89 healthy volunteers.

### Sample processing and sequencing

In this study, all samples were collected based on phase 2 protocol in the ImmuNexUT^27^; 27 immune cell types were purified from peripheral blood of each donor (**Supplementary Table 1)**. We first isolated PBMCs by density gradient separation with Ficoll-Paque (GE healthcare) immediately after the blood draw. Erythrocytes were lysed with Ammonium-Chloride-Potassium lysing buffer (Gibco), and non-specific binding was blocked with anti-human Fc-gamma receptor antibodies (Thermo Fisher Scientific). We next sorted PBMCs into 26 immune cell types with purity > 99% using a 14-color cell sorter BD FACSAria Fusion (BD Biosciences) with the aim of 5,000 cells per sample. The immune cell gating strategy for flow cytometry was based on the Human Immunology Project with slight modification^62^. Sorted cells were lysed and stored at −80°C. RNA was extracted using MagMAX-96 Total RNA Isolation Kits (Thermo Fisher Scientific). Libraries for RNA-seq were prepared using SMART-seq v4 Ultra Low Input RNA Kit (Takara Bio). Neutrophils were purified using MACSxpress Neutrophil Isolation Kits human (Miltenyi Biotec) with the aim of 2×10^6^ cells immediately after the blood draw, lysed and stored at −80°C, followed by RNA isolation with an RNeasy Mini Kits (QIAGEN) and library preparation with SMART-seq v4 Ultra Low Input RNA Kits (Takara Bio). All prepared libraries were sequenced on HiSeq2500 (6169 samples) or NovaSeq6000 (217 samples) (Illumina) to generate 100 or 150 base paired-end reads, respectively. Genomic DNA was isolated from peripheral blood using QIAmp DNA Blood Midi kit (QIAGEN). Libraries were prepared using TruSeq DNA PCR-Free Library prep kit (Illumina), followed by whole-genome sequencing (WGS). WGS was performed only for the samples from Japanese individuals. The details of WGS data processing were reported in our previous study^27^.

### Quantification and normalization of the expression data

Adaptor sequences were trimmed using Cutadapt (v1.16) and reads containing low-quality bases (Phred quality score < 20 in > 20% of the bases) were removed. Reads were aligned against the GRCh38 reference sequence using STAR^63^ (v2.5.3) with the UCSC (downloaded from illumina iGenome reference collection, archive-2015-08-14-08-18-15) and expression was counted with HTSeq^64^.

We applied multiple sample quality control (QC) steps to ensure high quality data. The samples with uniquely mapped read rates < 80% or unique read counts < 5 × 10^6^ were excluded as low-quality samples. To exclude outlier samples, we calculated Spearman’s correlations of the expressions between two samples from the same cell type and then removed the samples with mean correlation coefficients < 0.9. In addition, to exclude potentially swapped samples, we calculated the concordance rates between RNA-seq-based genotype and WGS-based genotype at the heterozygous loci and excluded samples with concordance rate < 0.9.

We then filtered out low expression genes (<10 counts or <1 count per million [CPM] in > 85% of samples), followed by a trimmed mean of M values (TMM) normalization with R (v4.0.2) package edgeR (v3.32.1)^65^ in each cell type. Normalized expression data were converted to log-transformed count per million (i.e., log[CPM+1]). The batch effects (i.e., product lots in SMART-seq v4 and sequencer; **Extended Data Fig. 2a**) were removed using Combat software^66^. To verify the successful work of the batch correction procedure, we used principal variance component analysis; we first calculated the explained variance of each clinical parameter for each PC1-7 score with the linear mixed models in R package lme4 (v1.1-27.1)^67^ and then inferred the average value of the explained variance weighted by each PC’s eigenvalue (**Extended Data Fig. 2b**).

### PCA and UMAP of all samples

For PCA and UMAP using all samples (**Extended Data Fig. 1b-c**), we combined the expression data after batch correction from each cell type and used the intersection of the genes (n=8397) that passed the filtering of low expression in each cell type. For UMAP, we used R package uwot (v 0.1.10) with default parameters^68^.

### PCA in each cell type of the discovery dataset

For PCA in each cell type of the discovery cohort, we used the top 10,000 variable genes from the expression data after batch correction in each cell type (**Fig. 2b; Extended Data Fig. 3**).

To calculate the proportion of sum squared deviations within HC, SLE, and between HC and SLE data in each cell type (**Fig. 2c**), we used the PC1-7 data of the discovery dataset (**Supplementary Note**). We first calculated the proportion of sum squared deviations for each PC score and then inferred the average value of the proportion weighted by each PC’s eigenvalue.

### Weighted variance partitioning analysis in each cell type

To calculate the explained variance of each clinical parameter within SLE transcriptome data for each cell type, we performed weighted variance partitioning analysis using the PC1-7 data of the discovery dataset (**Supplementary Note**). We first calculated the explained variance of each clinical parameter for each PC score using the linear mixed models in R package variancePartition (v1.20.0)^69^ and then inferred the average value of the explained variance weighted by each PC’s eigenvalue (**Fig. 2d, 5c; Extended Data Fig. 8, 9c; Supplementary Fig. 2b**).

To verify whether the inferred explained variance was not biased by outlier samples, we estimated standard errors (S.E.) of the explained variance by jackknife resampling method. When we had *n* samples for one cell type, we re-calculated the explained variance *n* times by excluding each one of the samples. We then evaluated the distribution of *n* explained variance and quantified its S.E. For each clinical parameter, we compared the explained variance in each cell type against the median explained variance across all 27 cell types. To assess the significance of the difference observed in this comparison for a cell type, we utilized *n* explained variance calculated in jackknife resampling for that cell type; among *n* values, we calculated the proportion of the values which was smaller than the median value, and we defined this proportion as jackknife resampling *P* (*P_jk_*). For each clinical parameter, if one cell type passed the Bonferroni-corrected *P_jk_* < 0.05, we concluded that the clinical parameter significantly contributed to the within-SLE transcriptome variation in that cell type.

### Linear models for the association between PC scores and clinical parameters

This study focused on the clinical parameters related to disease-state, overall disease-activity, organ/domain activity, and treatment statuses (**Supplementary Fig. 1a**). Disease-state was defined as the contrast between inactive SLE (i.e., not all SLE) and HC in the discovery dataset to exclude the elements of disease-activity signatures from the case-control contrast (**Fig. 1, middle**). For overall disease activity, we defined four categories: i) inactive as SLEDAI-2K^28^ = 0, ii) low disease activity (LDA) as 1 ≤ SLEDAI-2K ≤ 4, iii) moderate disease activity (MDA) as 5 ≤ SLEDAI-2K ≤ 8, and iv) high disease activity (HDA) as SLEDAI-2K ≥ 9. For organ activity, we categorized the patients into seven groups based on their actively involved organ/domains of the British Isles Lupus Assessment Group (BILAG) 2004^29^ and SELDAI-2K: a) constitutional, b) mucocutaneous, c) musculoskeletal, d) renal, e) extrarenal severe (neuropsychiatric/eye, cardiorespiratory and/or gastrointestinal), f) hematological, and g) serological activities. We also evaluated the effect of therapeutic agents such as MMF, hydroxychloroquine (HCQ), and tacrolimus (TAC)^3,60,70^.

To examine the associations between the PC scores (PC1-30) and clinical traits, we fitted the PC scores to the following linear regression models:

1. For disease-state (*x*: inactive SLE vs. HC),

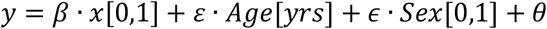

Here, *y* represents the scaled PC score for each cell type (PC1-30 × 27 cell types). All PC scores were scaled across samples to enable the direct comparison of the effect sizes in the associations with clinical parameters. In this comparison, age and sex were included as covariates.
2. For disease-activity (*x*: HDA vs. inactive SLE; we also examined LDA vs. inactive and MDA vs. inactive SLE),

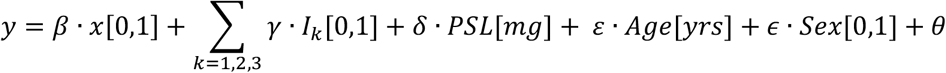

Here, *I*_*k*_ (*k* = 1,2,3) represents each immunosuppressant (MMF, HCQ, TAC) as covariates.
3. For organ/domain activity (*x_j_*[*j* = 1 … 7]: the abovementioned seven categories),

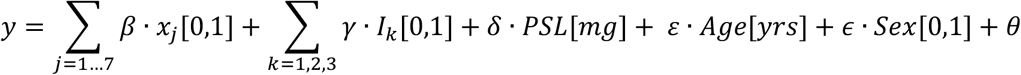

Here, we constructed multiple linear regression models including all seven categories, which enabled us to infer the association of each organ activity with PC scores, controlling the other organs’ effects (**Fig. 5c-d**).
4. For therapeutic agents (*I*_*k*_ [*k* = 1,2,3]: MMF, HCQ, TAC),

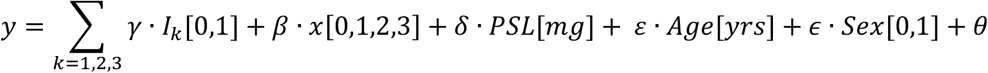

Here, *x* represents disease activity (inactive, LDA, MDA, and HDA) as covariates.

These equations enabled us to derive the associations of disease-state, activity, organ involvements or treatment statuses with PC scores, adjusted for other confounding factors. Statistical significance was set at FDR < 0.05. As described in **Supplementary Note** and **Supplementary Fig. 1b**, most of the significant associations were detected within PC1-7, with larger numbers than average per PCs. Therefore, we confirmed that PC1-7 is a minimum set to associate the transcriptome with the clinical parameters in the discovery dataset and utilized PC1-7 scores for the subsequent analyses. All PC scores were signed so that the effect sizes of disease-state and disease-activity were positive (**Fig. 5a-b; Extended Data Fig. 7a-b**). In the case of some PCs that had opposite sign in the effect sizes of disease-state and activity, the association with lower *P* value was prioritized to have positive effect size.

In the hierarchical clustering of 225 unique individuals using 189 PCs (= 7 PCs × 27 cell types) in the discovery dataset, the Euclidean distances of the PC scores were used with Ward’s method (**Fig. 5a; Extended Data Fig. 7a**).

### Differential gene expression analysis

To detect DEGs in each cell type, we fitted the TMM-normalized counts in the discovery dataset to the generalized linear models (GLM) with negative binomial distribution using edgeR (v3.32.1)^65^. The equations in these GLM models were consistent with those in the linear models as described in the **Methods**; **Linear models for the association between PC scores and clinical parameters**; we utilized the equations (1), (2) and (4). Additionally, we also considered the batch effects as covariates in this analysis since TMM-normalized counts were not corrected for batch effects (**Methods**; **Quantification and normalization of the expression data**). These equations enabled us to derive DEGs related to our focused disease traits or treatment statuses, adjusted for other confounding factors (**Fig. 3a-b, 6e; Supplementary Fig. 2a**). Statistical significance was set at FDR < 0.05. We defined (1) “disease-state signature genes” as significant DEGs between inactive SLE and HC, and (2) “disease-activity signature genes” as significant DEGs between HDA and inactive SLE for each cell type (**Fig. 3a**).

To evaluate the similarities and differences between disease-state and activity signature genes, we calculated Jaccard similarity indexes as the ratio of the shared genes with the concordant sign between the disease-state and activity signatures (**Fig. 3a**; orange dots) over the union of these two signature genes (**Fig. 3a**; orange + red + blue dots) for each cell type. Considering the biological significance, we did not regard the DEGs with the discordant sign between these two signatures as shared genes. Based on the proportion of DEGs and Jaccard index, we classified 27 cell types into three patterns (**Fig. 3b**). We first defined the cell types with Jaccard index > 0.15 as shared pattern and then classified the other cell types into disease-state or disease-activity dominant patterns based on which signature genes were numerically predominant.

We also calculated the Jaccard similarity index and Spearman correlation across all pairs of different cell types for both signature genes. The Jaccard similarity distances (i.e., 1 -Jaccard similarity indexes) of each pair within disease-state signature genes were used for hierarchical clustering (**Fig. 3d**). Similarly, Spearman’s correlation distance of each pair within disease-activity signature genes were used for hierarchical clustering (**Extended Data Fig. 4d**).

To detect DEGs related to therapeutic agents adjusted for confounding factors (e.g., disease activity), we set the patients not taking the agent as the control, meaning the down-DEGs represented the genes that were downregulated by each agent. Therefore, we calculated Jaccard indexes as the ratio of the shared genes with the inverse sign between the disease-activity signatures and MMF-DEGs over the union of these two signature genes (**Fig. 6e**). We calculated Jaccard indexes within the cell types in which more than 300 MMF-DEGs were observed.

### Replication analysis

For replication analysis, we compared our data with the three external bulk immune cell RNA-seq dataset of SLE and/or HC from different ancestries.

i. Cohort 1 (Panwar *et al.*^33^)

- Samples: 64 SLE and 24 HC, multi-ancestry cohort (Caucasian, Asian, Hispanic, and African)
- Cell subsets: six cell types (bulk T cells, bulk B cells, CL Mono, mDC, pDC, and Neu). Only CL Mono was collected from all donors, and the other five cell types were collected from around 20 SLE and 10 HC samples.
- Data usage strategy: since this cohort included both SLE and HC, we used this cohort for the replication analysis of disease-state and activity signatures (**Extended Data Fig. 5a-c**). Naive CD4 and naive B data from the current study were compared with bulk T and bulk B data from cohort 1, respectively (**Supplementary Table 4**).
ii. Cohort 2 (Takeshima *et al.*^35^)

- Samples: 30 SLE and 37 HC, All East Asian (EAS)
- Cell subsets: 19 cell types (Naive CD4, Mem CD4, Th1, Th2, Th17, Tfh, Fr. II eTreg, Naive CD8, bulk memory CD8 [Mem CD8], NK, Naive B, USM B, SM B, DN B, plasmablast, CL Mono, CD16p Mono, mDC, and pDC)
- Data usage strategy: this is our previous cohort independent of the ImmuNexUT. In this study, we excluded the overlapped samples with the current study, leaving relatively stable 30 SLE patients for the analysis. Therefore, we used this cohort only for the replication of disease-state signatures (**Extended Data Fig. 5a, c**). EM CD8 data from the current study was compared with bulk Mem CD8 data from cohort 2 (**Supplementary Table 4**).
iii. Cohort 3 (Andreoletti *et al.*^34^)

- Samples: 57 White and 63 Asian SLE patients
- Cell subsets: four cell types (bulk CD4 cells, NK cells, bulk B cells, and bulk monocytes)
- Data usage strategy: since this cohort did not include HC samples, we used this cohort only for the replication of disease-activity signatures (**Extended Data Fig. 5b-c**). Naive CD4, naive B, and CL Mono data from the current study were compared with bulk CD4, bulk B, and bulk monocyte data from cohort 3, respectively (**Supplementary Table 4**).

In all replications, we assessed the concordance of the directions of disease-state and activity signature genes in the discovery cohort with the corresponding genes in external cohorts using one-sided binomial sign tests (**Extended Data Fig. 5a-b**). The definitions of clinical status (e.g., disease-state and activity) were consistent with our discovery cohort, with the exception that inactive SLE in cohort 2 was defined as 0 ≤ SLEDAI-2K ≤ 2 (10 patients) since there were no patients with SLEDAI-2K = 0. In all replication analyses, we adjusted for the covariates (e.g., age and sex) in line with the analyses of the discovery dataset where applicable.

In the PC projection method, we first computed the Z score matrix of gene expressions using the mean and standard deviation (SD) of the discovery dataset and then inferred the PC scores of each sample from the external datasets as the inner products of each PC loading (**Supplementary Data 1**) and the Z score matrix. We tested the association of these PC scores with disease-state (i.e., inactive SLE vs. HC) and disease-activity (i.e., HDA vs. inactive SLE) in the external datasets using the linear regression model as with the discovery dataset (**Supplementary Table 9**). We then assessed the concordance of the directions of the effect sizes for disease-state and disease-activity PCs, respectively, using one-sided binomial sign tests (**Supplementary Note; Extended Data Fig. 5c**).

### Transcription factor and pathway enrichment analysis

To test pathway and transcription factor (TF) enrichment in each disease-state and activity signature genes, we performed over-representation analyses with one-sided Fisher’s exact test in R package clusterProfiler (v.3.18.1)^71^. Statistical significance was set at FDR < 0.05. For TF datasets, we used the Molecular Signatures Database (MsigDB) C3 all TF targets annotation (1133 annotations)^72^. For pathway datasets, we used the MsigDB hallmark gene set collection (50 annotations)^73^ and Kyoto Encyclopedia of Genes and Genomes (KEGG) pathway (548 annotations)^74^. To capture the cell-type-specific biology linked to disease-state and activity signatures genes, we set the union of both signature genes in all cell types as the background gene sets. For treatment-related DEGs (e.g., MMF-DEGs and BLM-DEGs), we performed pathway enrichment analysis only in the cell types in which more than 300 DEGs were observed.

### Analysis of pre- and post-BLM dataset

All 22 individuals received BLM treatment according to the standard protocols^6,7,75^. In this section, we defined those whose original disease-activity categories were moved into one or more lower categories (e.g., MDA to LDA or LDA to inactive) between pre- and post-BLM treatment, as good responders, and the others as poor responders.

Because edgeR did not implement generalized linear mixed models (GLMM), we detected DEGs between pre- and post-BLM using the following GLMM with negative binomial distribution in lme4 (v1.1-27.1)^67^, setting the statistical significance at FDR < 0.05 (**Fig. 6a-b**). Of note, we need to consider the batch effects as covariates in the following equation, since TMM-normalized counts were not corrected for batch effects (**Methods**; **Quantification and normalization of the expression data**).

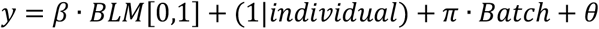

Here, *y* and *BLM* represents the TMM-normalized count for each gene in each cell type and the treatment status of BLM (i.e., pre- and post-BLM). We included a term for random intercept of individuals, and hence excluded individual-specific covariates (e.g., age and sex). In this equation, we set pre-BLM as the control, meaning the down-DEGs represented the genes that were downregulated by BLM treatment. Therefore, we calculated Jaccard indexes as the ratio of the shared genes with the inverse sign between the disease-activity signatures and BLM-related DEGs over the union of these two signature genes (**Fig. 6b**). We calculated Jaccard indexes within the cell types in which more than 300 BLM-DEGs were observed. Moreover, in each cell type, we compared associations between the logFC of disease-activity effect and those in BLM effect using linear regression model (**Supplementary Note; Extended Data Fig. 9a-b**).

In the PC projection method, we first computed the Z score matrix of gene expressions using the mean and SD of the discovery dataset and then inferred the PC scores of duplicated samples as the inner products of each PC loading (**Supplementary Data 1**) and the Z score matrix. To test the association between the change of PC scores and the treatment status, we used the following linear mixed model:

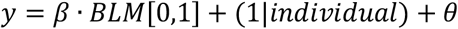

Here, *y* represents the scaled PC score for each cell type. In this model, it was not necessary to consider the batch effects since the PCA was performed using the log(CPM+1) gene expression data that had been already corrected for batch effects (**Methods**; **Quantification and normalization of the expression data**).

### Stratified linkage disequilibrium score regression

To evaluate the enrichment of the genome-wide distribution of all SLE risk variants irrespective of their effect sizes (heritability) around HC-SEG, disease-state and activity signature genes, we performed S-LDSC^26,54^. We examined the enrichment of SLE heritability for common variants within 100-kb windows on either side of the transcription start site of the genes with the top 1,000 highest Z-scores in either signature genes for each cell type, adjusting for baseline model provided by the developers^54^ (**Fig. 7a**). For this analysis, we used two large-scale SLE GWAS summary statistics from EAS^23^ and EUR ancestries^22^ (**Extended Data Fig. 10a**). Since the regression coefficients of S-LDSC are influenced by the GWAS heritability, we normalized coefficients by dividing them with mean per-SNP heritability as reported by a previous report^26^; we then reported normalized coefficients. In a fixed-effect meta-analysis of the two results, we used the inverse variance weighting method using normalized coefficients and their S.E. We reported *P* values to test whether the regression coefficient is significantly positive.

To call HC-SEG (specifically expressed genes in HC) for each cell type, we compared the expression data of one cell type with that of the remaining cell types that belong to other cell lineages using the GLM with negative binomial distribution in edgeR (v3.32.1)^65^. To be in line with previous studies of S-LDSC, only the samples from HC were used in this analysis.

### GWAS candidate genes enrichment analysis

The SLE-GWAS results were downloaded from the NHGRI-EBI GWAS Catalog^76^ on 16/08/2021. Among them, we defined the genes nearest to SLE-GWAS significant variants (*P* < 5×10^-8^) as GWAS candidate genes (**Supplementary Table 14**). Gene symbols were based on UCSC definition. To test the enrichment of GWAS candidate genes for disease-state and activity signature genes, we performed over-representation analyses with one-sided Fisher’s exact test (**Fig. 7b**). We set the union of the genes that passed the filtering of low expression in each cell type and used it as the background.

### Integrative analysis with eQTL data

To compare the direction between the risk allele’s expression quantitative trait loci (eQTL) effects and disease-state and activity signature genes, we utilized the results of the colocalization test between SLE-GWAS and ImmuNexUT eQTL data reported in Ota *et al*^27^. For visualization, logFC sign information was adjusted so that the direction of the coherent genes, which showed the concordant direction between eQTL effects for risk alleles and differential expressions^56^ (**Fig. 7e**), was positive (i.e., adjusted logFC).

To examine the interactive effects of the signature genes on the eQTL effects of SLE risk variants, we fitted the eGene expressions to the following linear regression models for each cell type (**Fig. 7f**):

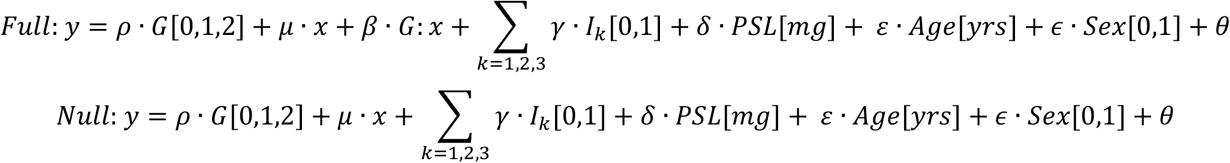

Here, *y* and *x* represents the expression of eGene and pGene, respectively, and *G* represents the genotype of each individual. *I_k_*(*k* = 1,2,3) represents each immunosuppressant (MMF, HCQ, TAC) as covariates. We tested the significance of interaction terms (i.e., *G*: *x*) by comparing full and null models using analysis of variance (ANOVA). Statistical significance was set at FDR < 0.05.

### Data availability

All analysis results including DEG list and PC loading scores are available as supplementary table and data. RNA-seq data used in this study will be available at the National Bioscience Database Center (NBDC) Human Database (Dataset ID: JGAS000486) upon acceptance.

### Code availability

Codes utilized in this study are available on GitHub (https://github.com/MasahiroNakano-hub).

## Data Availability

All analysis results including DEG list and PC loading scores are available as supplementary tables and data. RNA-seq data used in this study will be available at the National Bioscience Database Center (NBDC) Human Database upon publication.

## Acknowledgments

The super-computing resource was provided by Human Genome Center, Institute of Medical Sciences, The University of Tokyo (http://sc.hgc.jp/shirokane.html). This study was supported by Chugai Pharmaceutical Co., Ltd., Tokyo, Japan, the Ministry of Education, Culture, Sports, and the Japan Agency for Medical Research and Development (AMED) (JP21tm0424221 and JP21zf0127004). We appreciate all SLE patients and HC volunteers who participated in this study. We would like to thank all collaborators in the University of Tokyo, National Center for Global Health and Medicine, St. Luke’s International Hospital, and Chugai Pharmaceutical Co., Ltd for the contribution to sample collection and processing. M.N. and K.I. thank Michihiro Kono for helpful feedback. H. Suetsugu is supported by Practical Research Project for Rare/Intractable Diseases from Japan AMED. X.Y. is supported by Distinguished Young Scholar of Provincial Natural Science Foundation of Anhui (1808085J08). S. Bae is supported by Basic Science Research Program through the National Research Foundation of Korea funded by the Ministry of Education (NRF-2021R1A6A1A03038899).

## Author contributions

M.N. and K.I. conceived and designed the study. M.N. and K.I. wrote the manuscript with critical inputs from M.O. and K.F. M.N. conducted all analyses with the help of K.I. M.N., M.O., Y.T., Y.I., H.H., Y.N., T.I., J.M., R.Y., S.Y., A.N., Haruka T., Hideyuki T., Y.A., T.K., and H. Shoda managed and contributed to sample collection and processing. M.O., H.H., Y.N., and T.I. contributed to QC of the RNA-seq data. H. Suetsugu, L.L., K.K., X.Y., S. Bang, Y.C., H.L., X.Z., S. Bae, and C.T. managed and contributed to sample collection of EAS SLE-GWAS data. K.Y., T.O., and K.F. designed and managed the project. All authors contributed to the final version of the manuscript.

---

**Extended Data Fig. 1.**
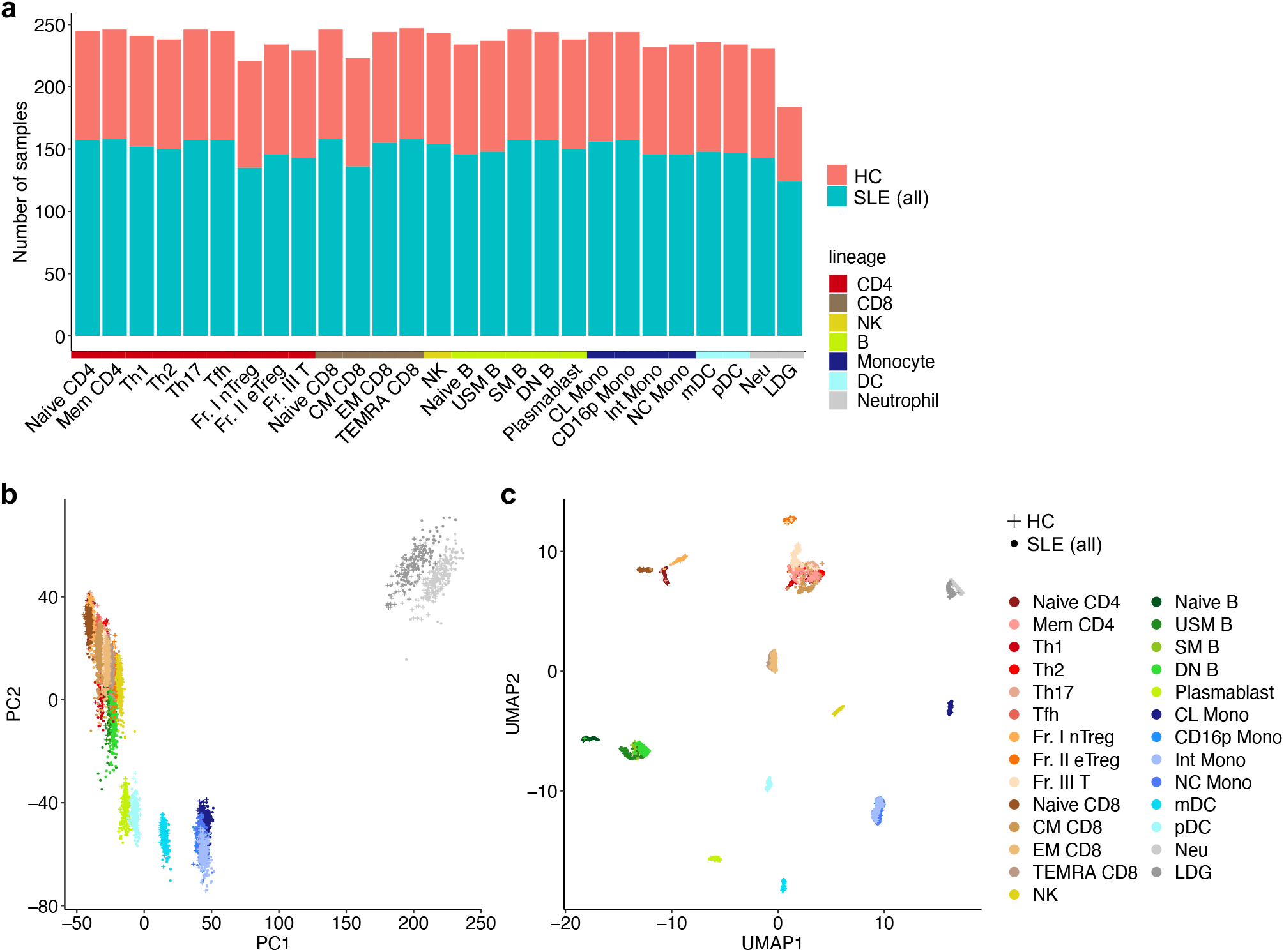
Overview of gene expression patterns in the ImmuNexUT cohort. **a,** Bar plots showing the number of samples that passed quality control (QC, **Methods**) in each cell type. **b,** A PCA and **c,** a UMAP plot of all samples. Colors and shapes indicate cell types and diseases, respectively. We used all 6,386 samples from 248 donors for all analyses in this figure.

**Extended Data Fig. 2.**
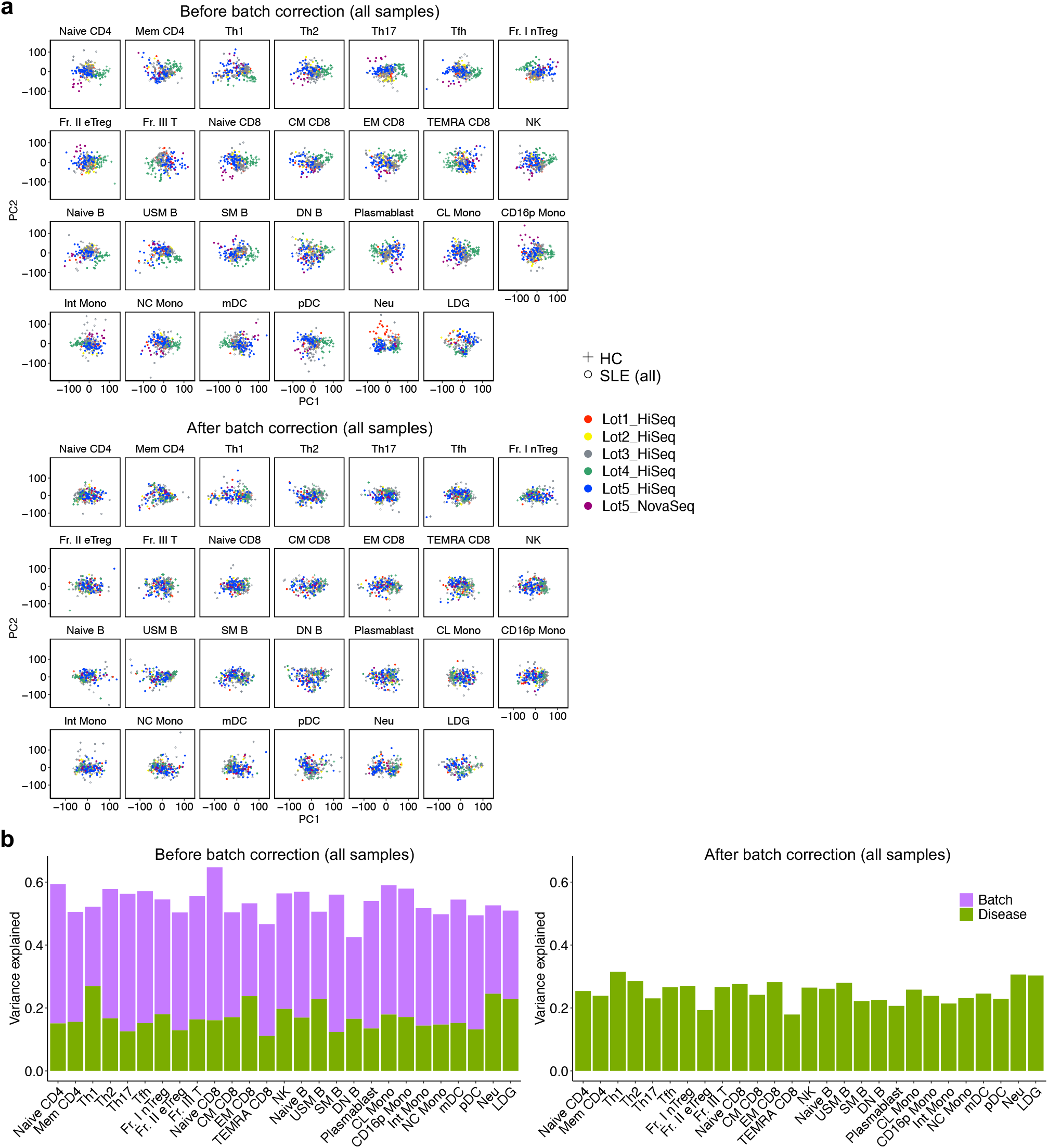
Batch correction procedure in this study. **a,** PCA plots of HC and all SLE gene expression data in each cell type **(top)** before and **(bottom)** after batch correction. Colors and shapes represent each batch and disease, respectively. **b,** Bar plots showing the proportion of variance explained by batch effect and disease in the gene expression data for each cell type **(left)** before and **(right)** after batch correction. We used all 6,386 samples from 248 donors for all analyses in this figure.

**Extended Data Fig. 3.**
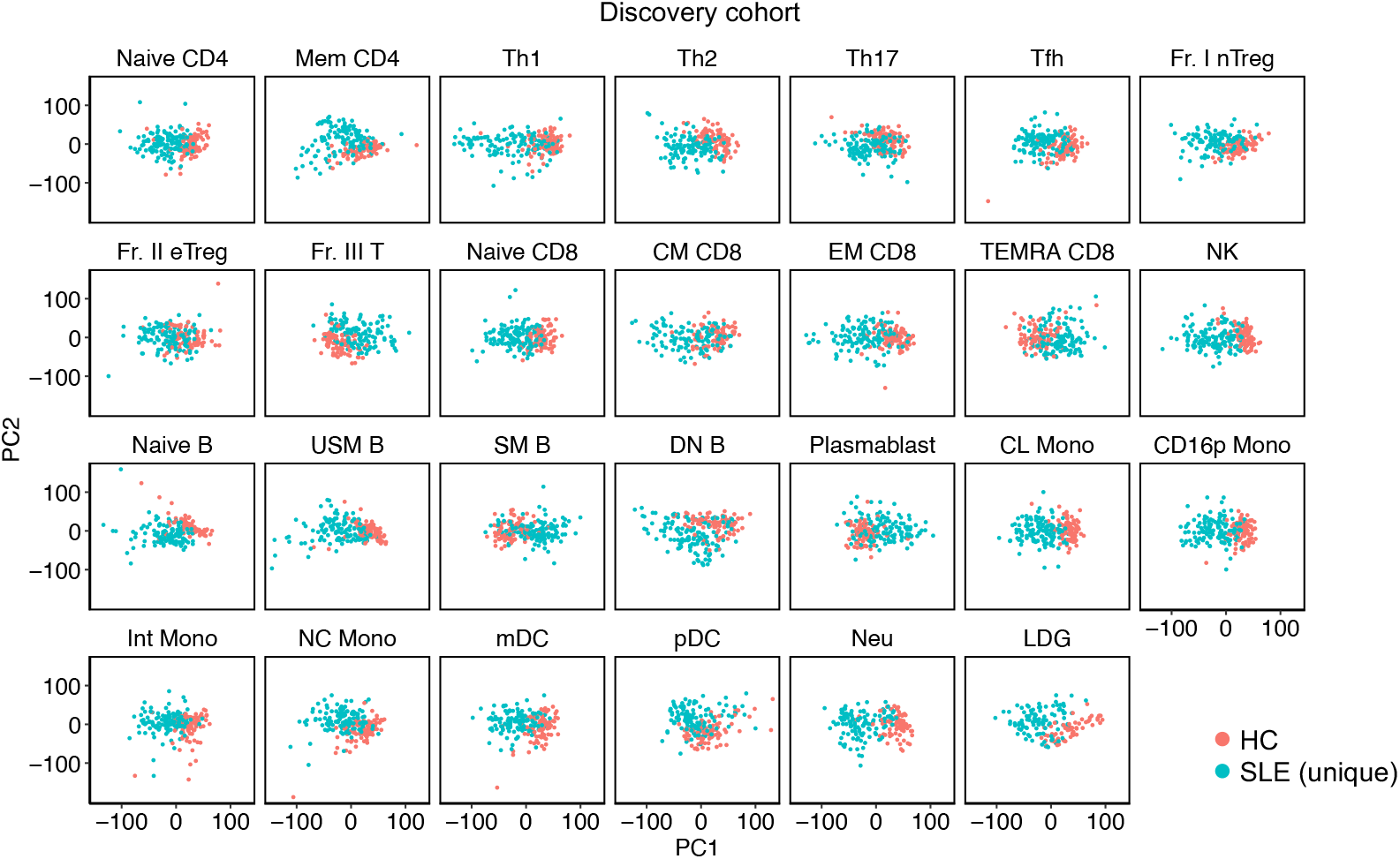
Overview of gene expression patterns in the ImmuNexUT cohort. PCA plots of HC and unique SLE gene expression data in each cell type. We used the discovery dataset (n=225) for this analysis.

**Extended Data Fig. 4.**
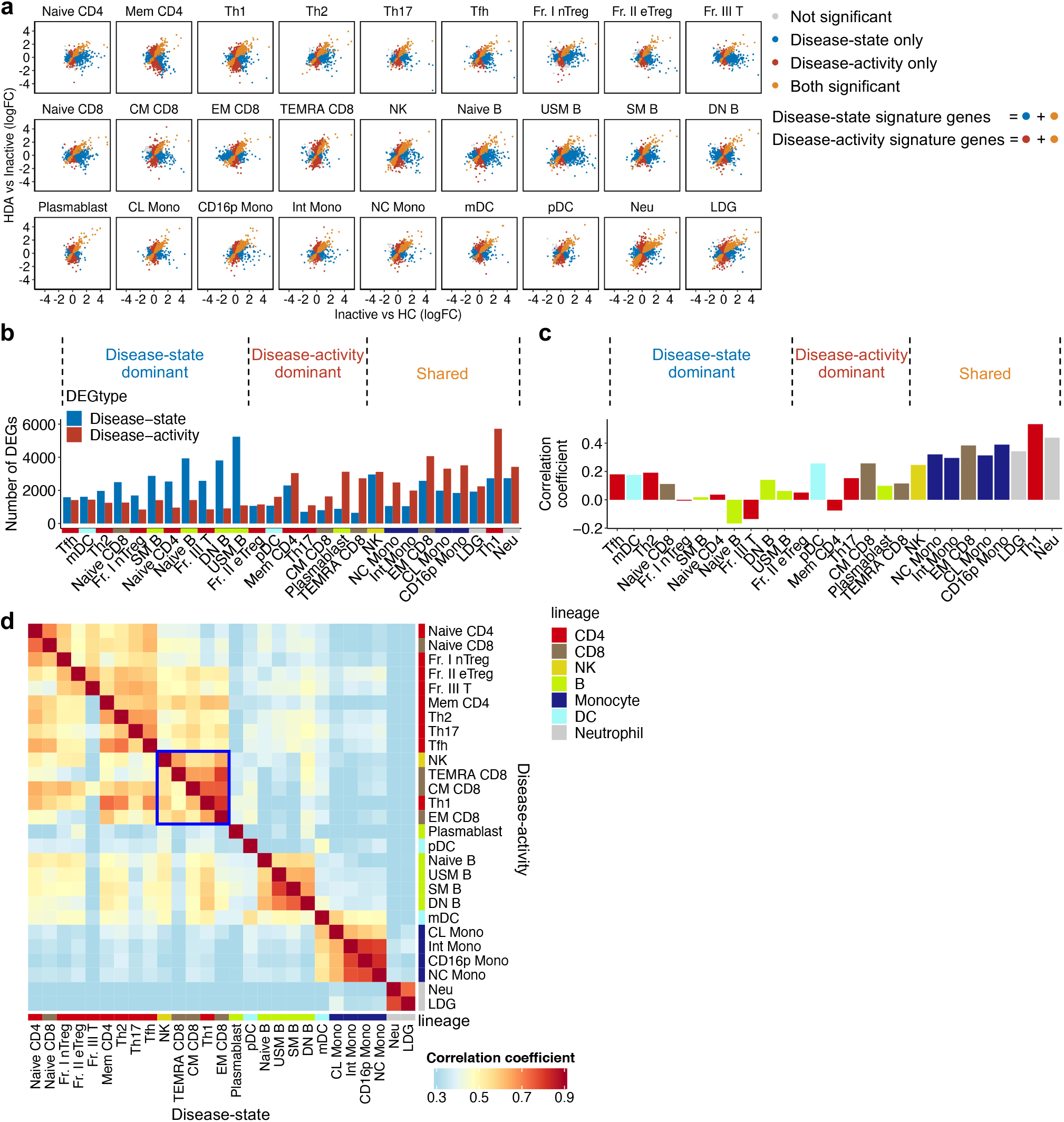
SLE disease-state and activity signatures. **a,** Scatter plots comparing the logFC of disease-state and activity signature genes in all cell types. Each dot represents one gene, colored based on the significance (FDR < 0.05 in differential expression test) of each comparison. Genes with logFC > 5 are plotted at the position of logFC = 5. **b,** A barplot showing the numbers of disease-state and activity signature genes in each cell type. **c,** A barplot showing the Spearman correlations between the logFC in disease-state and activity signatures for each cell type. In **b** and **c**, cell types are separated into three groups (**Methods**). **d,** A heatmap showing the Spearman correlations across all cell types in disease-state and activity signatures. The order of cell types in row and column are same and based on the hierarchical clustering using the Spearman correlation coefficients of disease-activity signatures. In **b** and **d**, row and column annotation colors indicate cell lineages. We used the discovery dataset (n=225) for all analyses in this figure.

**Extended Data Fig. 5.**
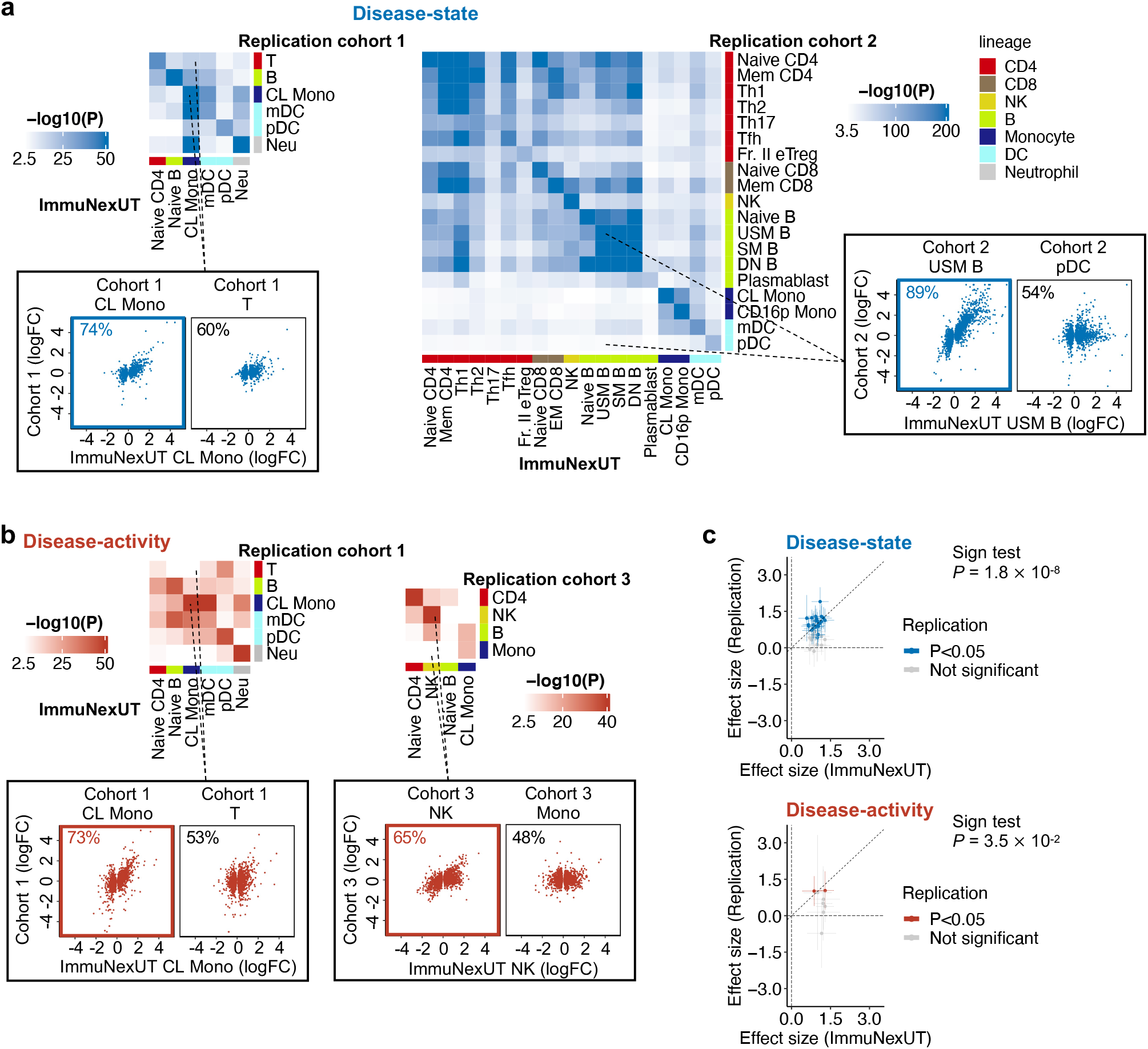
Lupus disease-state and activity signatures are replicable in independent cohorts. **a, (top)** Heatmaps showing the concordance between the disease-state signatures in the current study (HC, n=89; inactive SLE, n=31) and those in replication cohort 1 (left heatmap; HC, n=24; inactive SLE, n=16) and 2 (right heatmap; HC, n=37; inactive SLE, n=10). **(bottom)** Scatter plots comparing disease-state effects (logFC) of the current study and those of replication cohort 1 (left panel) and 2 (right panel) in representative corresponding (left plot in blue frame) and non-corresponding (right plot in black frame) cell-type combinations. **b, (top)** Heatmaps showing the concordance between the disease-activity signatures in the current study (inactive, n=31; HDA SLE, n=30) and those in replication cohort 1 (left heatmap; inactive, n=16; HDA SLE, n=6) and 3 (right heatmap; inactive, n=41; HDA SLE, n=4). **(bottom)** Scatter plots comparing disease-activity effects (logFC) of the current study and those of replication cohort 1 (left panel) and 3 (right panel) in representative corresponding (left plot in red frame) and non-corresponding (right plot in black frame) cell-type combinations. *P*, *P* values in one-sided sign test. In all heatmaps, only the combinations that pass Bonferroni-significance are colored. In scatter plots, each dot represents one signature gene. Genes with logFC > 5 in either comparison are plotted at the position of logFC = 5. **c, (top)** Scatter plots comparing the effect sizes of disease-state PCs in the current study and those in replication cohorts (cohort 1 and 2). The PCs with nominal *P* < 0.05 in linear regression test in replication cohorts are colored. **(bottom)** Scatter plots comparing the effect sizes of disease-activity PCs in the current study and those in replication cohorts (cohort 1 and 3). The PCs with nominal *P* < 0.05 in linear regression test in replication cohorts are colored.

**Extended Data Fig. 6.**
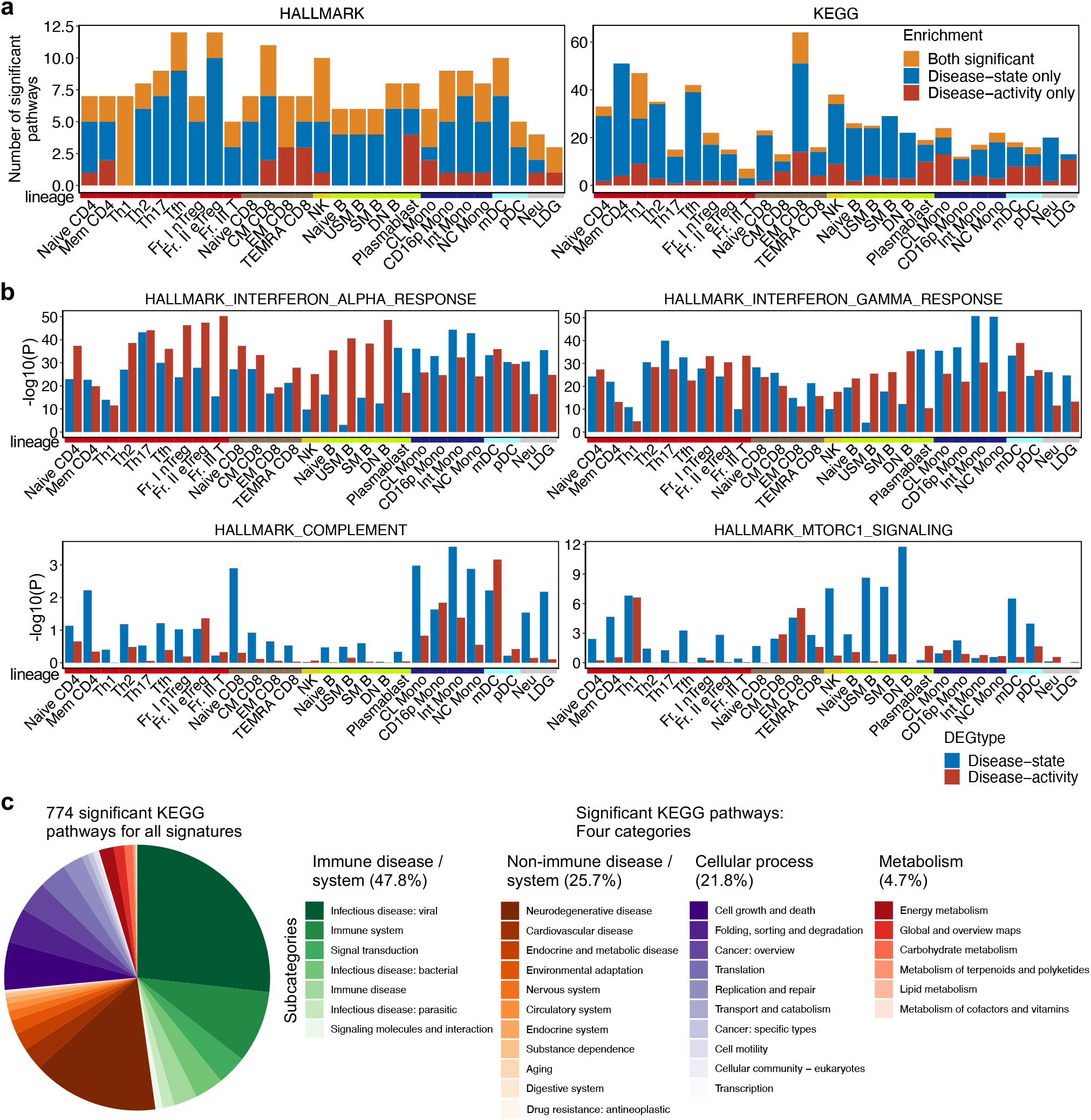
Cell-type-specific biology in disease establishment and exacerbation. **a,** Bar plots showing the number of significant **(left)** MsigDB HALLMARK and **(right)** KEGG pathway enrichments for disease-state and activity signature genes in each cell type. **b,** Bar plots showing the enrichment of representative MsigDB HALLMARK pathways for each signature. **c,** Classification of 774 KEGG pathway annotations significantly enriched in any signatures (FDR < 0.05 in one-sided Fisher’s exact test) into four main categories and 33 subcategories (**Methods**). We used the discovery dataset (n=225) for all analyses in this figure.

**Extended Data Fig. 7.**
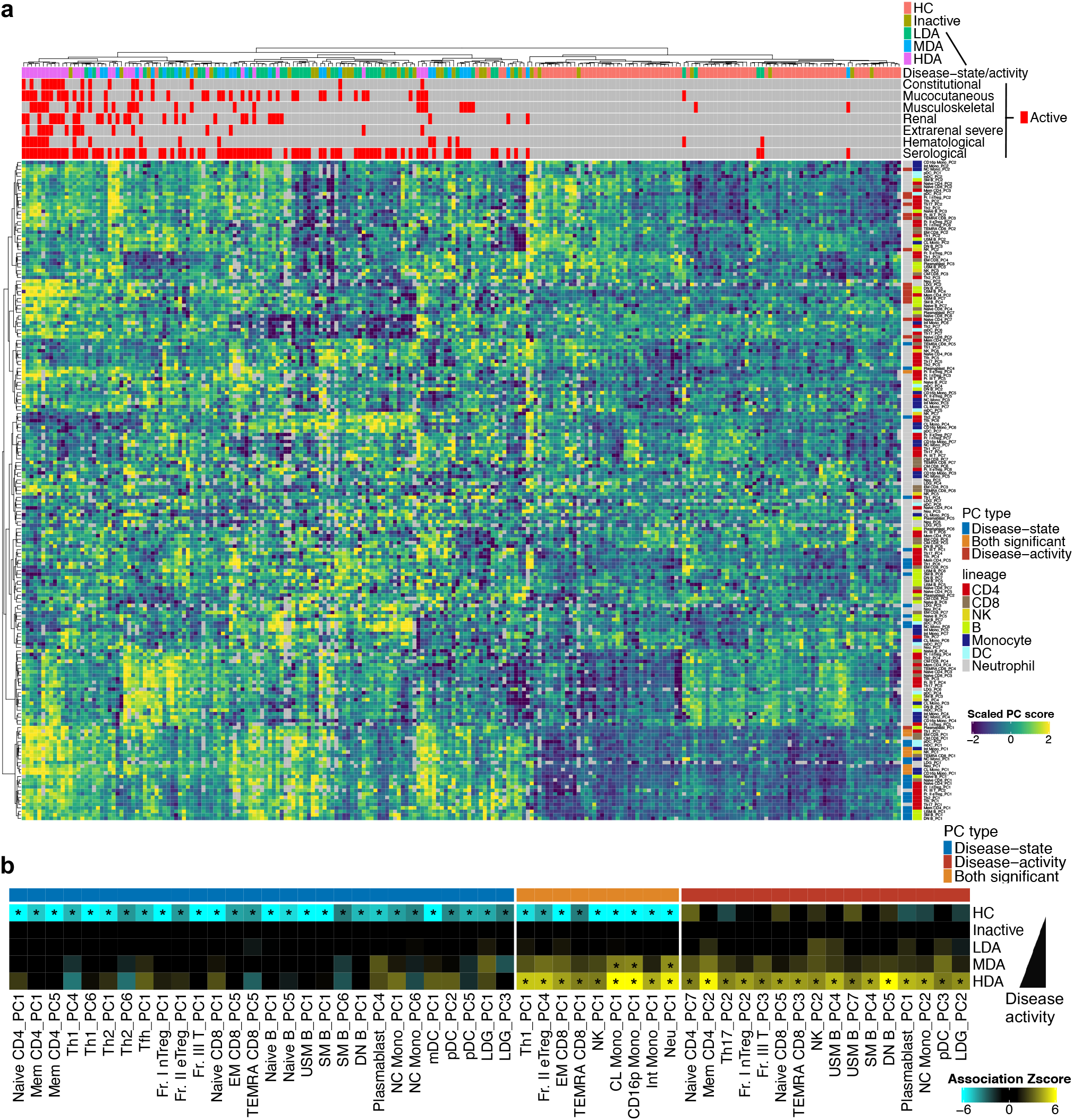
Disease-state and activity PCs in the discovery dataset. **a,** Hierarchical clustering of 225 unique individuals based on all PC1-7 scores of 27 cell types. Top annotations indicate the disease status and organ/domain activities in each individual. Right annotations indicate the type and cell lineage of each PC. **b,** A heatmap showing the association of PC scores with disease-state/activity in linear regression test. All PCs with significant association with disease-state and/or disease-activity are shown (*, FDR < 0.05). We used the discovery dataset (n=225) for all analyses in this figure.

**Extended Data Fig. 8.**
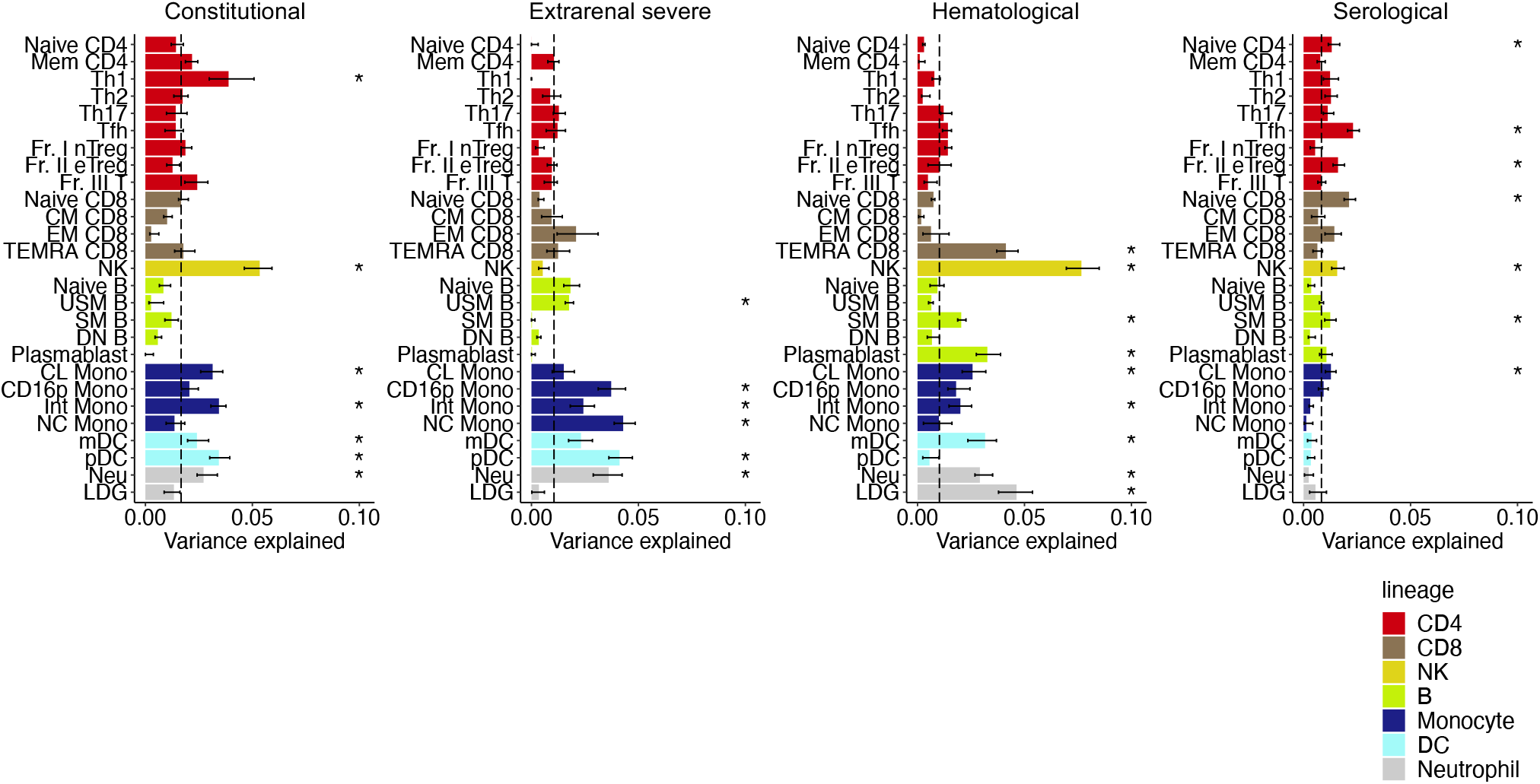
Cell-type-specific contribution to organ involvement in SLE. Bar plots showing the proportion of variance explained by organ/domain activities within SLE data in each cell type. Here, the results for the four organ/domain activities other than those in Fig. 5c**, right** are shown. Error bars and dashed vertical lines indicate 95% confidence intervals from jackknife resampling and the median values across 27 cell types, respectively. *, Bonferroni-adjusted *P*jk < 0.05 (**Methods**). We used SLE patients in the discovery dataset (n=136) for this analysis.

**Extended Data Fig. 9.**
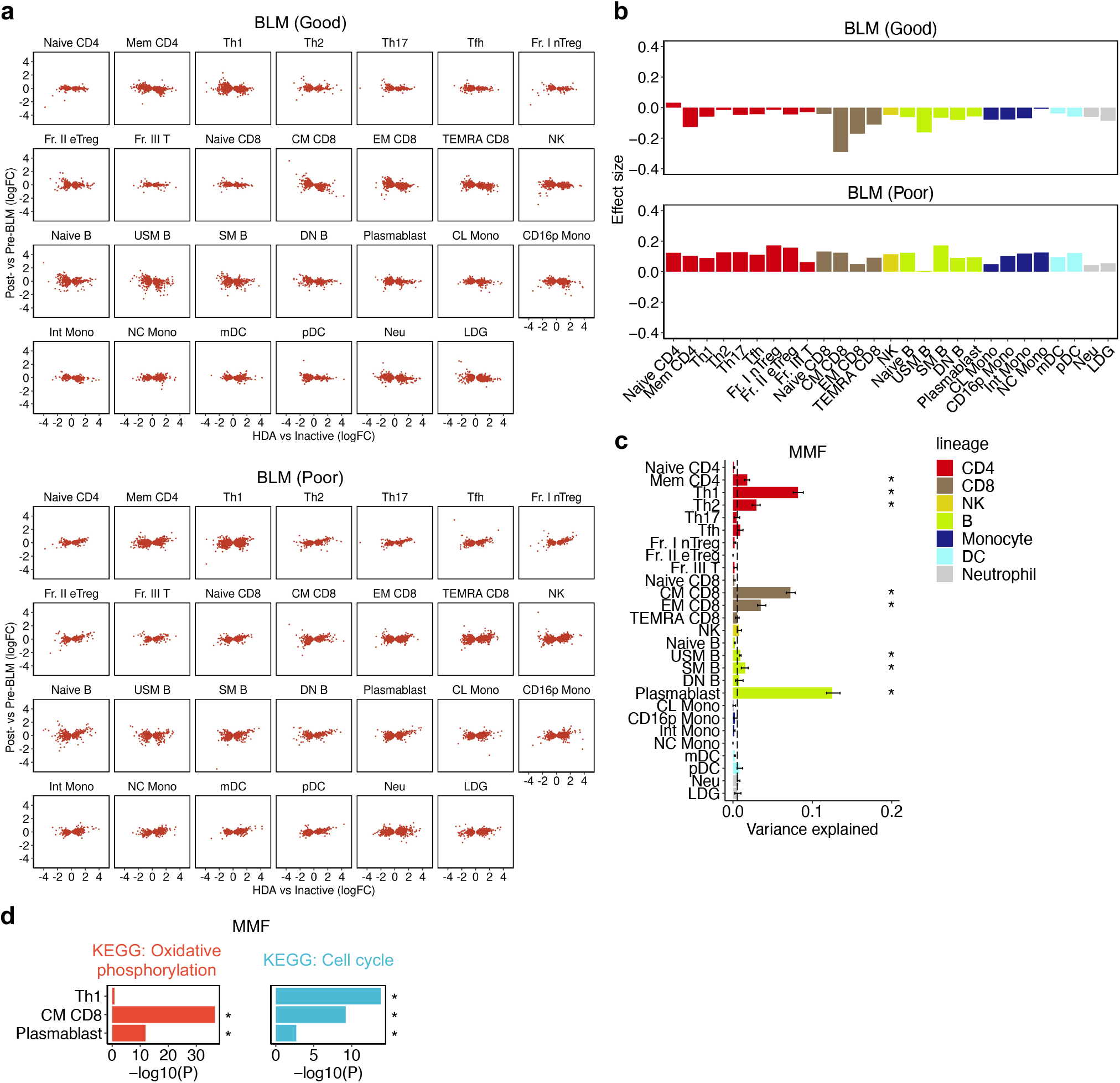
Cell-type-specific activity signatures linked to treatment responses. **a,** Scatter plots comparing the disease-activity effects (logFC) and BLM effects (logFC) of the activity signature genes in each cell type, separated into good and poor responders. Each dot represents one gene. Genes with logFC > 5 are plotted at the position of logFC = 5. **b,** Bar plots showing the effect sizes in the linear regression tests for the association between disease-activity effects and BLM effects, separated into good and poor responders. **c,** Bar plots showing the proportion of variance explained by medication status of MMF within SLE data in each cell type. Error bars and dashed vertical lines indicate 95% confidence intervals from jackknife resampling and the median values across 27 cell types, respectively. *, Bonferroni-adjusted *P*jk < 0.05 (**Methods**). **d,** Bar plots showing the enrichment of representative pathways for the MMF-DEGs in Th1, CM CD8, and plasmablast. *P*, nominal *P* values; *, FDR < 0.05 in one-sided Fisher’s exact test. For BLM-related analyses, we used the pre- (n=22) and post-BLM (n=22) patients. For MMF-related analyses, we used the patients with (n=31) and without (n=105) MMF.

**Extended Data Fig. 10.**
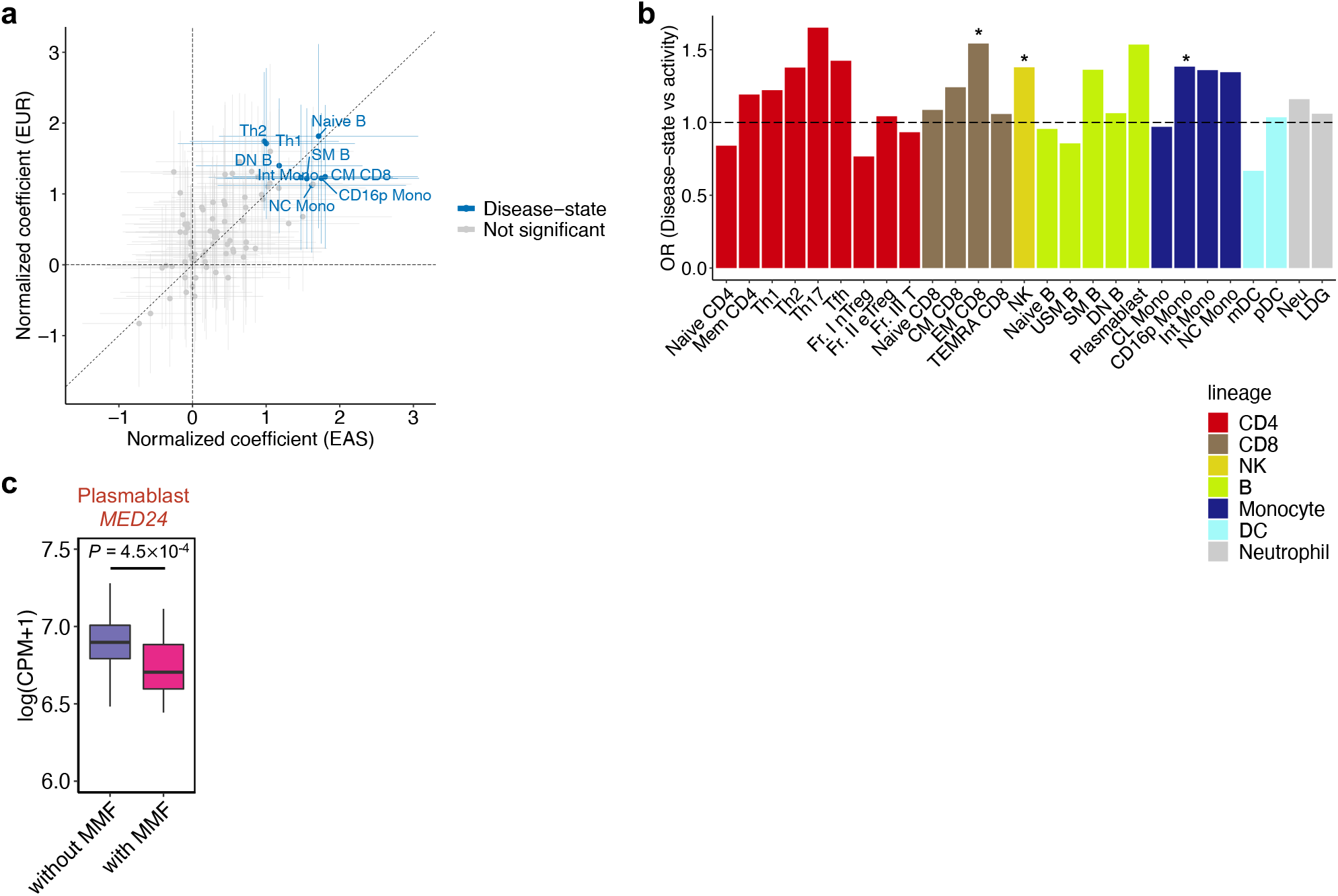
Risks variants for SLE are enriched around disease-state signatures, not activity signatures. **a,** Scatter plots comparing the normalized coefficients in S-LDSC for EAS and EUR SLE-GWAS. Each dot represents each signature (i.e., [HC-SEGs, disease-state and disease-activity signatures] × 27 cell types). The signatures that pass Bonferroni-significance in meta-analysis are annotated. **b,** A bar plot showing the direct comparison of GWAS candidate genes enrichment between disease-state and activity signature genes for each cell type using one-sided Fisher’s exact test. Dashed horizontal line indicates odds ratio (OR) = 1. *; nominal *P* < 0.05. **c,** A Box plot showing the expression of *MED24* in plasmablasts between patients with and without MMF. *P*, *P* values in differential expression test. Within each boxplot, the horizontal lines reflect the median, the top and bottom of each box reflect the IQR, and the whiskers reflect the maximum and minimum values within each grouping no further than 1.5 x IQR from the hinge. We used the discovery dataset (n=225) for all analyses in this figure.

