## Supplementary material for "Cell-type-specific transcriptome architecture underlying the establishment and exacerbation of systemic lupus erythematosus": Nakano_SLE_transcriptome_Sup_final.pdf

### Supplementary Note

#### Study design differences between our study and previous studies.

Compared with previous studies with fine resolution transcriptomes[1–3], larger sample size with multiple clinical statuses (e.g., disease activity, organ involvement, and treatment profiles) is an advantage of our cohort (**Supplementary Table 2**). For example, while a PBMC scRNA-seq study of SLE recruited only seven and three inactive and HDA patients in the main dataset, respectively[3], we recruited 31 and 30 inactive and HDA patients, respectively. While recent tissue scRNA-seq studies only focused on ~24 kidney and ~17 skin samples from SLE patients[1,2], we included a total of 43 other organ involvements (e.g., musculoskeletal, neurological, cardiorespiratory, and gastrointestinal) in addition to 30 renal and 41 mucocutaneous activities. Furthermore, our cohort also included 22 pairs of pre- and post-BLM treatment data.

Although scRNA-seq would be the mainstream of future transcriptome studies, the strategy that we took in this bulk, but fine resolution transcriptome study has complemented the limitations of recent scRNA-seq studies of SLE[1–3], e.g., smaller sample size and sparse expression information at a single cell level. As a result, we successfully profiled transcriptome features of multiple immune cells in a variety of patient conditions.

#### Strategies to select PC in this study.

To evaluate how the transcriptome variation reflected the heterogeneous clinical statuses in the discovery dataset, we next tested their associations with the PC scores of each cell type using a linear regression model, adjusted for potential confounding factors (e.g., age, sex, and treatments; **Methods**). Since cumulative explained variance reached 50-60% until PC30 in all cell types, we tested the association of 14 clinical parameters and PC1-30 scores for each cell type (**Supplementary Fig. 1a**). We identified 105 significant associations between the clinical parameters and the PC1-30 from all cell types (FDR < 0.05; **Supplementary Table 3**). Overall, most of the significant associations were detected within PC1-7, with larger numbers than average per PCs. (**Supplementary Fig. 1b**). Consistent with this observation, UMAP using all PC1-7 scores clearly distinguished clinically active SLE patients from HC (**Supplementary Fig. 1c**). Therefore, we confirmed that PC1-7 is a minimum set to associate the transcriptome with the clinical parameters in the discovery dataset and utilized PC1-7 scores for the subsequent analyses.

The primary aim of this study was to understand clinical heterogeneity of SLE from transcriptome perspective. Since transcriptome is affected by multiple factors (e.g., age, sex and genetic background) in addition to clinical parameters, the associations between the clinical parameters and the PCs could have been observed in the lower-ranked PCs. Therefore, the fact that top PCs dominantly reflected clinical heterogeneity indicates the success of our transcriptome study.

**Lupus disease-state and activity signature genes are highly replicable in independent cohorts.**
To assess the validity and generalizability of disease-state and activity signatures, we conducted replication analysis. We compared both signature genes with the immune cell RNA-seq data from the following three independent cohorts of SLE and/or HC from different ancestries (**Methods**). Importantly, the high cellular resolution is a big advantage of our discovery cohort, but at the same time it put hurdle on the replication analysis since many of the previous studies do not have the same-level cellular resolution as our study; hence this replication study is limited by imperfect cell-type matching (**Supplementary Table 4**).

i) Cohort 1 (Panwar *et al.*[4])

- Samples: 64 SLE and 24 HC, multi-ancestry cohort (Caucasian, Asian, Hispanic, and African)
- Cell subsets: six cell types (bulk T cells, bulk B cells, CL Mono, mDC, pDC, and Neu). Only CL Mono was collected from all donors, and the other five cell types were collected from around 20 SLE and 10 HC samples.
- Data usage strategy: since this cohort included both SLE and HC, we used this cohort for the replication analysis of disease-state and activity signatures. Naive CD4 and naive B data from the

current study were compared with bulk T and bulk B data from cohort 1, respectively (**Supplementary Table 4**).

ii) Cohort 2 (Takeshima *et al.*[5])

- Samples: 30 SLE and 37 HC, All East Asian (EAS)
- Cell subsets: 19 cell types (Naive CD4, Mem CD4, Th1, Th2, Th17, Tfh, Fr. II eTreg, Naive CD8, bulk memory CD8 [Mem CD8], NK, Naive B, USM B, SM B, DN B, plasmablast, CL Mono, CD16p Mono, mDC, and pDC)
- Data usage strategy: this is our previous cohort independent of the ImmuNexUT. In this study, we excluded the overlapped samples with the current study, leaving relatively stable 30 SLE patients for the analysis. Therefore, we used this cohort only for the replication of disease-state signatures. EM CD8 data from the current study was compared with bulk Mem CD8 data from cohort 2 (**Supplementary Table 4**).

iii) Cohort 3 (Andreoletti *et al.*[6])

- Samples: 57 White and 63 Asian SLE patients
- Cell subsets: four cell types (bulk CD4 cells, NK cells, bulk B cells, and bulk monocytes)
- Data usage strategy: since this cohort did not include HC samples, we used this cohort only for the replication of disease-activity signatures. Naive CD4, naive B, and CL Mono data from the current study were compared with bulk CD4, bulk B, and bulk monocyte data from cohort 3, respectively (**Supplementary Table 4**).

First, we tested the replicability of disease-state signatures using cohort 1 and 2. We confirmed high concordance in disease-state signatures (**Extended Data Fig. 5a**, one-sided sign test, $P < 10^{-34}$ and $P < 10^{-76}$ for each pair of corresponding cell types from cohort 1 and 2, respectively).

Next, we tested the replicability of activity signatures using cohort 1 and 3. Despite the limitations in the cell-type matching, we observed good concordance in activity signatures, supporting the reproducibility of our data (**Extended Data Fig. 5b**, one-sided sign test, $P < 10^{-8}$ and $P < 10^{-3}$ for each pair of corresponding cell types from cohort 1 and 3, respectively).

We thus demonstrated high reproducibility of our two categories of lupus-relevant signatures and their high transferability to other datasets, including more bulky gene expression data.

**Lupus disease-state and activity PCs showed a good replicability in independent cohorts.**

To assess the validity of disease-state and activity PCs in the discovery dataset, we projected the data from the three external datasets onto our PCA space in the corresponding cell types (**Methods; Supplementary Table 4**). We then tested the association of the inferred PC scores with disease-state (i.e., inactive SLE vs. HC) and disease-activity (i.e., HDA vs. inactive SLE) using the linear regression model (**Supplementary Table 9**). For disease-state PCs, 94% (33/35) of the PCs showed concordant sign (one-sided sign test, $P = 1.8 \times 10^{-8}$; **Extended Data Fig. 5c**). For disease-activity PCs, 88% (7/8) of the PCs showed concordant sign (one-sided sign test, $P = 3.5 \times 10^{-2}$). Thus, we successfully showed a good replicability of our two categories of lupus-relevant PC signatures.

**Pathway analysis using MsigDB hallmark gene set collection.**

We performed pathway enrichment analyses using 50 MsigDB hallmark gene set annotations (**Extended Data Fig. 6a, left; Supplementary Table 7**). We confirmed the strong enrichments of established hallmark pathways for SLE (**Extended Data Fig. 6b**):

i) IFN response in all cell types for both signatures[7–9]

ii) complement activation in monocyte-lineage cells and myeloid DC (mDC) for both signatures[10,11]

iii) mammalian target of rapamycin complex (mTORC) 1 signaling in lymphocytes and DC-lineage cells predominantly for disease-state signatures[12,13]

These findings indicate that our transcriptome data successfully recapitulated currently established SLE biology.

**The association of disease-activity PCs and organ activity.**

We evaluated the direct relationship of each disease-activity PC with organ involvement. Here, we constructed multiple linear regression models including all seven organ/domain categories, which enabled us to inter the association of each organ activity with PC scores, controlling the other organs' effects (**Methods**). As described in the main text, we identified four associations that passed FDR < 0.05: Neu PC1 and NC Mono PC2 for renal activity, and Naive CD4 PC7 and DN B PC5 for musculoskeletal activity (**Fig. 5d; Supplementary Table 3**). Additionally, we observed some interesting findings in other associations that did not reach statistical significance after multiple testing correction (23 associations that passed nominal $P < 0.01$): (i) CL Mono PC1 and musculoskeletal/renal activity, consistent with the variance partitioning analysis (**Fig. 5c**), (ii) USM B PC4 and extra-renal severe/serological activity, (iii) USM B PC7/SM B PC 4 for hematological activity. Thus, our results suggested potential cell-type-specific contributions to each organ involvement, which may be informative in unravelling SLE clinical heterogeneity.

**The association of BLM effects and disease-activity signature effects**

When we calculated the Jaccard similarity index between BLM-DEGs and activity signature genes, we restricted the analysis to B-lineage cells for which more than 300 BLM-DEGs were observed. To evaluate BLM effects on all cell types, we next extended this analysis to all activity signature genes, not restricting it to BLM-DEGs. We tested associations between logFC in disease-activity effects and those in BLM effects (**Extended Data Fig. 9a-b**). We observed that disease-activity signature effects, especially in B- and CD8+ memory T-lineage cells, were negatively correlated with the BLM effects only in good responders (linear regression test; FDR < 0.05).

### Supplementary Figures

**a**

| Disease trait | | Immunosuppressant |
|---|---|---|
| **Disease-state/activity** | **Organ/domain activity** | |
| Inactive vs. HC | Constitutional | HCQ |
| LDA vs. Inactive | Mucocutaneous | MMF |
| MDA vs. Inactive | Musculoskeletal | TAC |
| HDA vs. Inactive | Renal | |
| | Extrarenal severe* | |
| | Hematological | |
| | Serological | |

→ 14 clinical parameters × 27 cell types × PC1-30

*Extrarenal severe = Neuropsychiatric/eye + Cardiorespiratory + Gastrointestinal

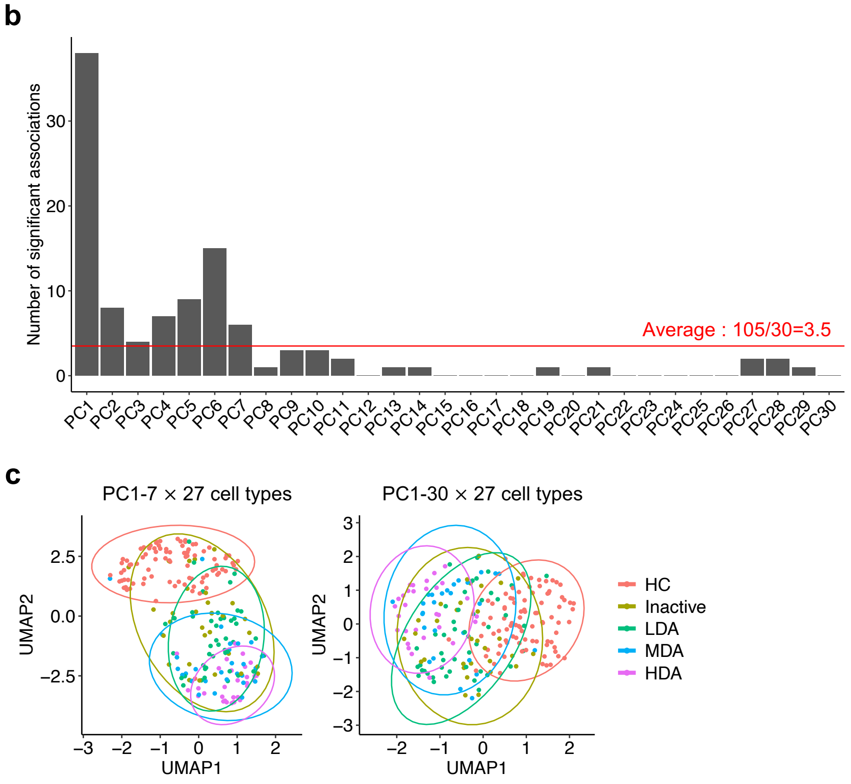


**Supplementary Fig. 1| Strategies to select PC in this study. a,** A list of the 14 clinical parameters examined in this study. The association of these parameters and the PC scores in each cell type are tested in linear regression model, adjusted for confounding factors **(Methods)**. **b,** A histogram showing the number of significant associations with the clinical parameters for the total 105 significant PCs in each cell type (FDR <0.05 in linear regression test). The red horizontal line indicates the average number of significant associations per one PC. **c,** A UMAP plot of 225 individuals in the discovery dataset using (**left**) PC1-7 and (**right**) PC1-30 scores in all 27 cell types. Colors represent the clinical status of individuals. LDA, low disease activity; MDA, moderate disease activity; HDA, high disease activity; HCQ, hydroxychloroquine; MMF, mycophenolate mofetil; TAC, tacrolimus. We used the discovery dataset (n=225) for all analyses in this figure.

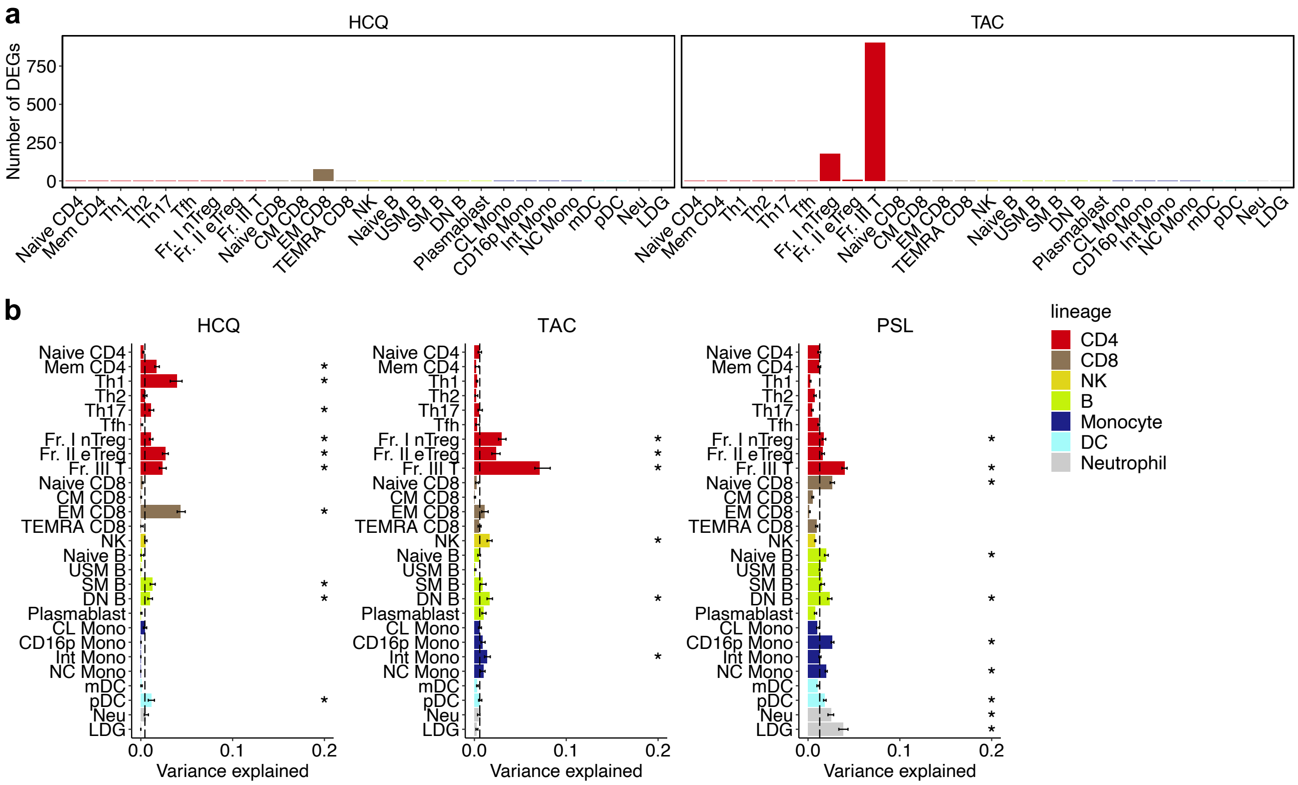


**Supplementary Fig. 2| Cell-type-specific activity signatures linked to treatment effect. a,** Bar plots showing the numbers of DEGs between patients with vs. without **(left)** hydroxychloroquine (HCQ) and **(right)** tacrolimus (TAC) in each cell type. **b,** Bar plots showing the proportion of variance explained by medication status of **(left)** HCQ, **(middle)** TAC, and **(right)** prednisolone (PSL) within SLE data in each cell type. Error bars and dashed vertical lines indicate 95% confidence intervals from jackknife resampling and the median values across 27 cell types, respectively. *, Bonferroni-adjusted $P_{jk}$ < 0.05 (**Methods**). We used SLE patients in the discovery dataset (n=136) for all analyses in this figure.
